# Systematic large-scale application of ClinGen InSiGHT *APC*-specific ACMG/AMP variant classification criteria substantially alleviates the burden of variants of uncertain significance in ClinVar and LOVD databases

**DOI:** 10.1101/2024.05.03.24306761

**Authors:** Xiaoyu Yin, Marcy Richardson, Andreas Laner, Xuemei Shi, Elisabet Ognedal, Valeria Vasta, Thomas v. O. Hansen, Marta Pineda, Deborah Ritter, Johan T. den Dunnen, Emadeldin Hassanin, Wencong Lyman Lin, Ester Borras, Karl Krahn, Margareta Nordling, Alexandra Martins, Khalid Mahmood, Emily A.W. Nadeau, Victoria Beshay, Carli Tops, Maurizio Genuardi, Tina Pesaran, Ian M. Frayling, Gabriel Capellá, Andrew Latchford, Sean V. Tavtigian, Carlo Maj, Sharon E. Plon, Marc S. Greenblatt, Finlay A. Macrae, Isabel Spier, Stefan Aretz

## Abstract

**Background:** Pathogenic constitutional *APC* variants underlie familial adenomatous polyposis, the most common hereditary gastrointestinal polyposis syndrome. To improve variant classification and resolve the interpretative challenges of variants of uncertain significance (VUS), APC-specific ACMG/AMP variant classification criteria were developed by the ClinGen-InSiGHT Hereditary Colorectal Cancer/Polyposis Variant Curation Expert Panel (VCEP).

**Methods:** A streamlined algorithm using the *APC*-specific criteria was developed and applied to assess all *APC* variants in ClinVar and the InSiGHT international reference *APC* LOVD variant database.

**Results:** A total of 10,228 unique *APC* variants were analysed. Among the ClinVar and LOVD variants with an initial classification of (Likely) Benign or (Likely) Pathogenic, 94% and 96% remained in their original categories, respectively. In contrast, 41% ClinVar and 61% LOVD VUS were reclassified into clinically actionable classes, the vast majority as (Likely) Benign. The total number of VUS was reduced by 37%. In 21 out of 36 (58%) promising *APC* variants that remained VUS despite evidence for pathogenicity, a data mining-driven work-up allowed their reclassification as (Likely) Pathogenic.

**Conclusions:** The application of *APC*-specific criteria substantially reduced the number of VUS in ClinVar and LOVD. The study also demonstrated the feasibility of a systematic approach to variant classification in large datasets, which might serve as a generalisable model for other gene-/disease-specific variant interpretation initiatives. It also allowed for the prioritization of VUS that will benefit from in-depth evidence collection. This subset of *APC* variants was approved by the VCEP and made publicly available through ClinVar and LOVD for widespread clinical use.

## INTRODUCTION

Familial adenomatous polyposis (FAP, OMIM #175100) is an autosomal dominant precancerous condition and the most common monogenic gastrointestinal polyposis syndrome, caused by constitutional (germline) pathogenic variants (PV) in the tumour suppressor gene *APC* (1–3). The colorectal phenotype exhibits high inter- and intra-familial variability from the growth of less than 100 up to thousands of adenomatous polyps (4). Surveillance colonoscopy and/or prophylactic (procto)colectomy are offered to prevent colorectal cancer or delay disease progression (5–7). Hence, the identification of an *APC* PV has direct relevance for the patients and their relatives, defining *APC* as a highly clinically actionable gene (8). *APC* is a large gene composed of 15 coding exons which encodes a multifunctional protein with several functional domains. Depending on the colorectal phenotype and family history, causative *APC* variants can be identified in up to 85% of patients with adenomatous polyposis (9–13), the vast majority of which are nonsense and frameshift variants leading to a truncated protein with abrogated function (5–7, 14). During the last three decades, thousands of rare *APC* PV have been identified in patients with FAP. Variants are distributed across the gene, the majority of which are private, observed in only one or very few families (www.lovd.nl/APC).

On the other hand, advances in high-throughput sequencing with the widespread implementation of large multi-gene panel testing (MGPT) and exome/genome sequencing have generated an additional plethora of *APC* variants in (healthy) individuals without a polyposis phenotype, many of which are missense alterations. In the absence of comprehensive data and consensus for the level of evidence required to corroborate variant interpretation, most of these variants remain Variants of Uncertain Significance (VUS) or variants with conflicting assertions, accounting for around 67% of *APC* variants in ClinVar. These VUS confer medical uncertainty and pose challenges in the clinical management of patients and their relatives.

Since its inception, the ACMG/AMP guideline has evolved through further refinements to the various variant assessment methods and evidence codes (15–21) and the development of gene- or disease-specific ACMG/AMP classification criteria by variant curation expert panels (VCEPs) under the governance of ClinGen (Clinical Genome Resource) (22). The International Society for Gastrointestinal Hereditary Tumours (InSiGHT) is a research consortium that houses and curates the world’s largest databases for variants of gastrointestinal cancer-predisposing genes in the Leiden Open Variation Database (LOVD) (23). Recently, a ClinGen-InSiGHT Hereditary Colorectal Cancer/Polyposis VCEP (HCCP VCEP) was established (https://clinicalgenome.org/affiliation/50099/). The *APC* subcommittee (*APC* VCEP) developed and validated APC-specific ACMG/AMP classification criteria (24), readying the VCEP for variant submissions to ClinVar as an FDA Recognized Expert Panel. The most updated version of the VCEP specifications can be found at https://cspec.genome.network/cspec/ui/svi/doc/GN089.

In this study, we used the *APC*-specific criteria to perform a large-scale reclassification exercise of all *APC* variants listed in ClinVar and the InSiGHT *APC* reference database LOVD. The criteria were embedded and applied in a streamlined algorithm, which was supplemented by further data mining and curation to achieve the most accurate classification. The results were compared to their original classifications in respective databases and any discrepancies or clinical conflicts were addressed.

## METHODS

### *APC* variant database merging and centralisation

Prior to the extraction of variants, the landscape of all publicly available databases containing *APC* variants was identified and examined for activity and curation status. Of at least 19 *APC* databases, nine are inactive and another three are not curated (Supplementary Table 1). All listed curators, in particular those of inactive, outdated, or orphaned databases were contacted to request sharing and merging of data with the reference LOVD (v.3.0) installation. To establish a centralised, curated data source of *APC* variants with consistent reporting format and phenotypic description, the InSiGHT *APC* LOVD (https://www.insight-database.org/genes/APC) and the Global Variome shared LOVD (www.lovd.nl/APC) were subsequently merged to generate one international reference *APC* gene variant database in LOVD, abbreviated in the following as LOVD and accessible via all three URLs.

### ClinVar and LOVD variant extraction and annotation

ClinVar variants with summary evaluation and individual submitter annotations were retrieved from the March 2022 XML file. All alleles associated with the *APC* gene were extracted. The merged *APC* LOVD database was downloaded on 12/05/2022. The legacy description of published variants was recorded alongside their standardised nomenclature as per the HGVS (Human Genome Variation Society) guidelines on the preferred reference transcript NM_000038.6 (25), correcting for any errors where possible. All non-structural variants were annotated using the Ensembl Variant Effect Predictor (VEP) (26). Structural variants defined by genomic alterations greater than 50bp in size (gross deletions, duplications, inversions, in-frame, Alu and SVA retrotransposon insertions, inversions and complex variants) were annotated manually using Mutalyzer (27).

### Reclassification algorithm

Details of the *APC-*specific criteria were as published previously (24). A stepwise algorithm encompassing all evidence codes was designed to systematically evaluate all *APC* variants in ClinVar and LOVD (Figure 1).

**Figure 1.**
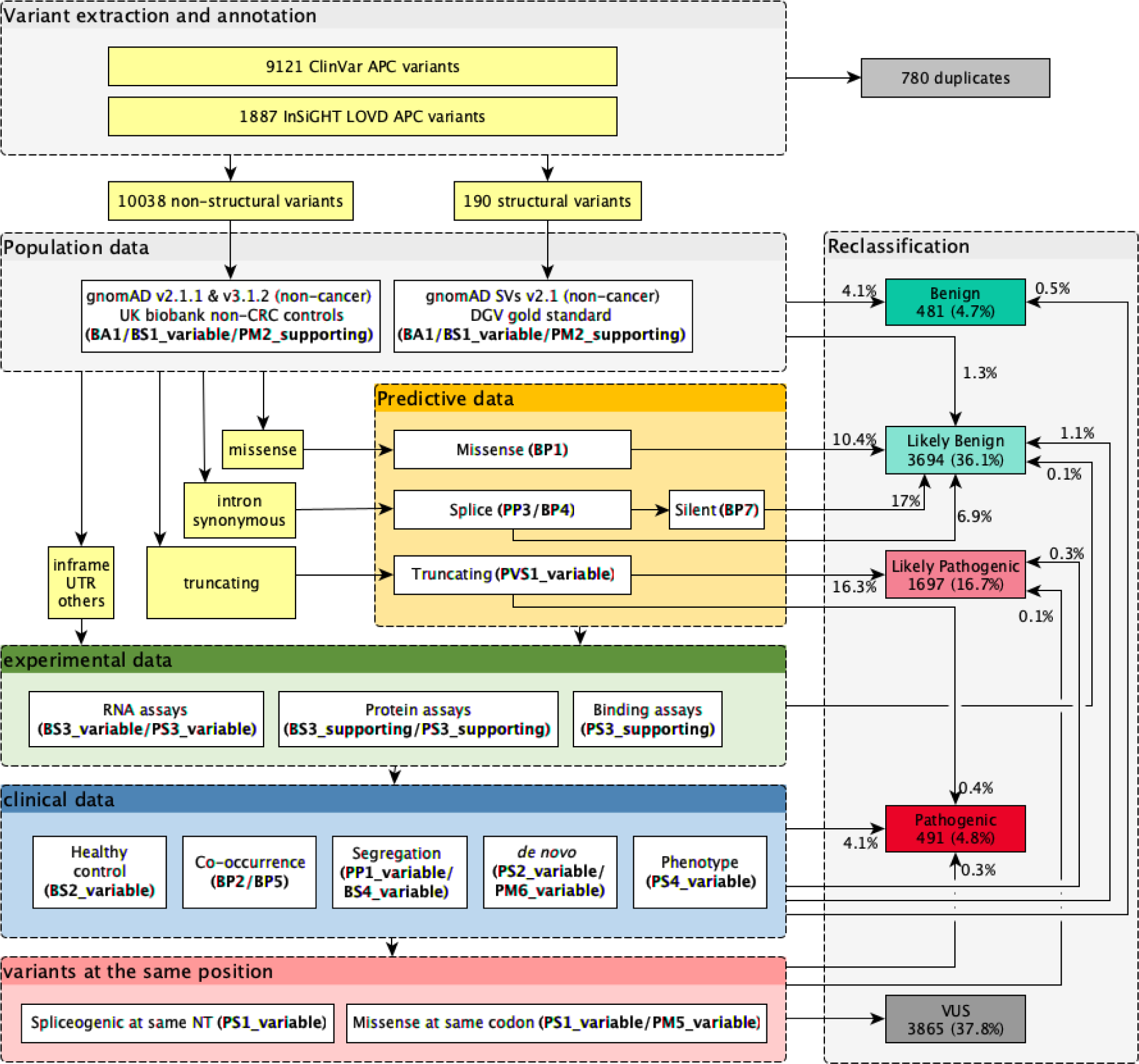
*APC-*specific criteria embedded in a reclassification algorithm. An algorithm demonstrating the application of all eligible *APC-*specific codes to *APC* variants in ClinVar and the InSiGHT LOVD) in a stepwise approach, and the percentage of variants that reached a B/LB or P/LP classification at each step. Firstly, the highest minor allele frequency of non-structural and structural variants was calculated from gnomAD non-cancer datasets or UK Biobank non-colorectal cancer control data, and gnomAD SVs (structural variants) or DGV (database of genomic variants) gold standard, respectively. Predictive criteria were then applied based on the most severe variant consequence as predicted by Ensemble VEP (variant effect predictor). Variants with any experimental and/or clinical evidence were identified and assigned corresponding code. Finally, splice variants at the same nucleotide and missense variants at the same codon were identified and the variants at the same position criteria were applied.

#### Minor allele frequency data (BA1, BS1, PM2_supporting)

The frequency of all *APC* variants in reference populations were compared against the minor allele frequency (MAF) criteria BA1 (≥ 0.001), BS1 (≥ 0.00001), and PM2_supporting (≤ 0.000003 or absent; first version of criteria). The non-cancer datasets from gnomAD (the Genome Aggregation Database) v2.1.1 and v3.1.2 were used as the reference population frequency data for non-structural variants (28). Exome sequencing data from 323,228 healthy individuals without a diagnosis of CRC in the UK Biobank were also used to further enhance the detection of rare APC variants (29). If a variant was present in multiple reference population datasets, the highest MAF was calculated from any subpopulation with more than 2000 alleles, with the exclusion of founder populations. The frequency of structural variants was examined in gnomAD SVs v2.1 and the DGV (Database of Genomic Variants) Gold Standard release from 15/05/2016 (30).

#### Predictive data (PVS1, PP3, BP1, BP4, BP7)

Truncating variants, canonical ± 1/2 splice site variants, and exonic last nucleotide guanine to non-guanine variants were assigned the Loss-of-Function (LoF) criterion PVS1 if they are located between codon 49 and 2645 inclusive (24). Splice prediction was performed using SpliceAI and MaxEntScan via VEP, which determined PP3 and BP4 eligibility for synonymous and intronic variants and PP3 eligibility for presumed missense variants to reveal possible splicing effects (31, 32). Variants exceeding a score of 0.6 in SpliceAI and a 15% reduction from the native site prediction in MaxEntScan were considered spliceogenic, and variants with a SpliceAI score of less than 0.2 and a MaxEntScan score of less than 3 were considered to have no impact on splicing. The criteria were only applied when both prediction tools showed concordant results. BP7 was subsequently applied to synonymous and deep intronic variants at or beyond +7/-21 which satisfied BP4. The missense code BP1 was applied to missense variants located outside of the first 15-amino acid repeat of the β-catenin binding domain (codon 1021-1035) if they were consistently deemed non-spliceogenic by SpliceAI and MaxEntScan.

#### Experimental data (BS3, PS3)

Published mRNA splicing assays and protein function assays of *APC* variants were collated in a systematic review by the *APC* VCEP to derive gene-specific recommendations for the application of the experimental criteria BS3 and PS3 (24). VCEP-approved experimental evidence for *APC* variant classifications included RNA assays for PS3 and BS3, β-catenin regulated transcriptional assays for PS3_supporting and BS3_supporting and surface plasmon resonance assays for PS3_supporting. The proportion of aberrant transcripts, evidence of biallelic expression and use of nonsense-mediated decay inhibition were also noted from the original publications when available, which determined the quality of the data and therefore the weight assigned for PS3 and BS3.

#### Clinical data (PS4, PS2, PM6, PP1, BS4, BP2, BP5, BS2)

The phenotype details of individuals with *APC* variants from the InSiGHT LOVD download were retrieved. In cases where phenotype details were recorded as unstructured text, text mining was employed to extract and stratify useful information. Affected individuals were scored for PS4 using methods as described previously (24). Further data mining was undertaken focusing on the identification of confirmed *de novo APC* variants (PS2/PM6), segregation and non-segregation analysis (PP1/BS4), co-occurrence of the variant under assessment with other established pathogenic *APC* variant (BP2) or with an alternative molecular basis of disease (BP5) including heterozygous PV in *POLD1* or *POLE* (Polymerase-proofreading-associated polyposis), and biallelic PV either in *MUTYH* (MUTYH-associated polyposis), in *NTHL1* (NTHL1-associated tumour syndrome), in *MSH3* (MSH3-associated polyposis), or the MMR genes *MLH1*, *MSH2*, *MSH6,* or *PMS2* (Constitutional MMR deficiency). The Human Gene Mutation Database (HGMD) and Universal Mutation Database (UMD) were also examined for additional data (33, 34). For variants that were already listed in gnomAD, their presence in healthy unaffected individuals in the UK Biobank non-CRC population was regarded as additional evidence in support of a benign classification (BS2). The absolute number of heterozygous individuals in the UK Biobank non-CRC dataset was counted, scored according to the *APC* VCEP’s definition for a healthy individual, and given the healthy control code BS2 with appropriate weight. As homozygous LoF *APC* variants were shown to be lethal at embryonic developmental stages (35), the observation of variant homozygosity ≥ 2 times in any reference population database was also considered strong evidence for a benign classification as defined in the *APC*-specific criteria.

### Review and synthesis of primary criteria combination

The *APC*-specific criteria applied for each variant were pulled together to generate a preliminary criteria combination according to the *APC* VCEP’s rules for combining criteria (24). In addition to the phenotype description in LOVD, for variants that fulfilled a reasonable set of predictive and/or experimental criteria (e.g., PVS1, PS3 or BS3) but remained unclassified, the corresponding internal clinical records and/or reference publication of the variant in question was consulted to curate additional phenotype points, and further data mining was conducted using Mastermind, LitVar and PubMed as required to reach a non-VUS classification. Additionally, the classification of the 58 published *APC* variants in the pilot study for the development of the *APC-*specific guideline were incorporated, adding another layer of data and expert review to the result. Finally, the revised criteria combination was used to calculate a final pathogenicity class. Missense variants at the same codon and spliceogenic variants at the same nucleotide position as established P or LP variants were identified and the criteria PS1 or PM5 were applied accordingly.

### Prioritised list of variants for further review

The revised variant classification was compared with the original pathogenicity assertion in ClinVar and LOVD. Variants with clinically significant conflict (B/LB vs. P/LP) between the original assertion and reclassification were examined for causes of discrepancy. From the variants that remained VUS after reclassification, 36 variants close to reaching a pathogenic classification were identified (Table 1 and Figure 2), which included (1) truncating VUS that fulfilled PVS1 but not PM2_supporting; (2) VUS with pathogenic *in silico* predictions; (3) VUS with experimental findings suggestive of deleterious effect and (4) VUS observed in individuals with FAP-associated phenotypes. A targeted literature search was conducted to acquire further information for all 36 variants. If a clinically relevant classification could be achieved, no further work-up was done (group 1 in Table 1 and Figure 2). If this was not the case, phenotype data were requested for these variants from VCEP members and internal laboratory contributors in a standardised clinical data request form and analysed with the aim to resolve conflicting criteria and upgrade classification (group 2 in Table 1 and Figure 2).

**Figure 2.**
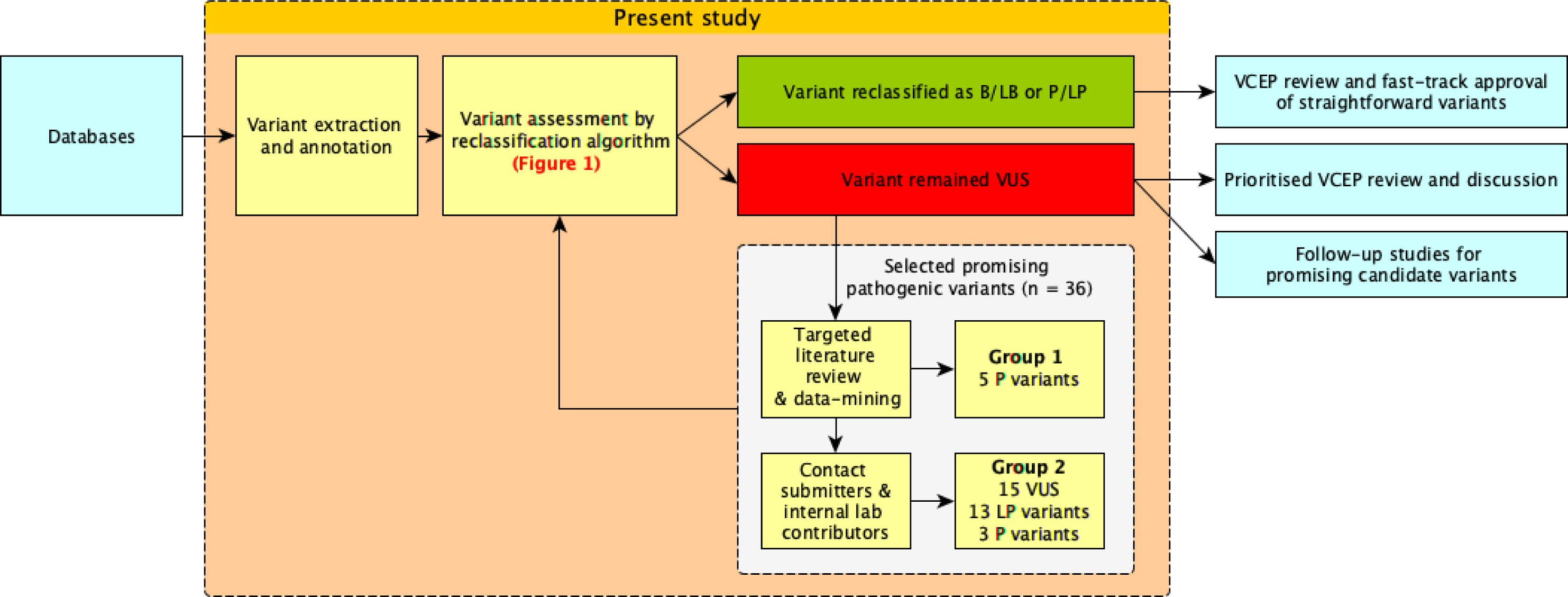
Reclassification workflow and suggested method of operation for ongoing VCEP activity. This workflow summarised the organisational procedures undertaken in this study for variant reclassification, validation of the reassigned class and prioritisation of variants with high clinical importance for different modes of VCEP review and approval. Relatively straightforward variants might be processed in batch and become candidates for fast-track VCEP approval (e. g. variants fulfilling BA1 or BS1 plus BP1). From the remaining VUS 36 variants with some evidence for pathogenicity were selected and five variants were re-assessed as Pathogenic by a targeted literature review and data mining (Group 1). The remaining 31 variants were reassessed based on further clinical information requested from respective ClinVar submitters and internal lab contributors, which ultimately leads to prioritised VCEP review.

**Table 1.** Further curation of selected variants remaining VUS after application of reclassification algorithm.

| HGVSc | Database_ID | Prior classification | Criteria applied by algorithm | Classification by algorithm | Further curation | Final criteria applied | Final classification of VCEP |
| --- | --- | --- | --- | --- | --- | --- | --- |
| **Group 1** Previously Pathogenic variants, for which further assessment based on literature review, data-mining and reassessment of minor allele frequency (MAF) criteria resulted in a classification of Pathogenic |  |  |  |  |  |  |  |
| c.471G>A (p.Trp157Ter) | ClinVar 411479 | Pathogenic | PVS1 | VUS | 1.5 phenotype points (47-50); revised MAF criteria | PVS1, PS4_supp, PM2_supp<sup>a</sup> | Pathogenic |
| c.1312+4_1312+19del | LOVD APC_001939 | Pathogenic | BP4, PM2_supp | VUS | 6.5 phenotype points, RNA assay result, segregated in 8 meiosis (51) | PS3_mod, PS4, PP1_strong, PM2_supp | Pathogenic |
| c.1333C>T (p.Gln445Ter) | ClinVar 438865 | Pathogenic | PVS1, BS1 | VUS | 1 phenotype point (52, 53); revised MAF criteria | PVS1, PS4_supp, PM2_supp<sup>a</sup> | Pathogenic |
| c.2546_2551del (p.Asp849_Ser851delinsGly) | LOVD APC_000075 | Pathogenic | PM2_supp, PS4_mod | VUS | Error in annotation: c.2546delATAGAAG (legacy name). Correct nomenclature: c.2546_2552del; p.(Asp849Valfs*10), 1 phenotype point, segregation in 3 meioses (11) | PVS1, PS4_supp, PM2_supp, PP1 | Pathogenic |
| c.4669_4670del (p.Ile1557Ter) | ClinVar 183857 | Pathogenic | PVS1 | VUS | 1 phenotype point (54, 55); revised MAF criteria | PVS1, PS4_supp, PM2_supp<sup>a</sup> | Pathogenic |
| **Group 2** Promising potentially pathogenic VUS, further assessment based on clinical evidence from ClinVar submitters and internal lab contributors and reassessment of MAF criteria |  |  |  |  |  |  |  |
| c.136-4A>G | 925741 | VUS | PP3, BS1 | VUS | observed in 1 individual worth 0 phenotype points; revised MAF criteria | PP3 | VUS |
| c.156del (p.Gly53GlufsTer17) | ClinVar 654864 | Pathogenic | PVS1 | VUS | observed in 1 individual worth 0 phenotype points; revised MAF criteria | PVS1, PM2_supp<sup>a</sup> | Likely pathogenic |
| c.203del (p.Leu68TyrfsTer2) | ClinVar 934724 | Pathogenic | PVS1 | VUS | observed in 4 individuals worth 1 phenotype point; revised MAF criteria | PVS1, PS4_supp, PM2_supp<sup>a</sup> | Pathogenic |
| c.220+2T>A | ClinVar 141515 | Likely pathogenic | PVS1, BS1 | VUS | observed in 35 individuals worth 8 phenotype points; revised MAF criteria | PVS1, PS4, PM2_supp<sup>a</sup> | Pathogenic |
| c.422G>C (p.Arg141Thr) | ClinVar 1056286 | VUS | PVS1_strong, PM2_supp | VUS | observed in 3 individuals worth 0 phenotype points | PVS1_strong, PM2_supp | VUS |
| c.422G>A (p.Arg141Lys) | ClinVar 824696 | VUS | PVS1_strong | VUS | observed in 0 individual worth 0 phenotype points; revised MAF criteria | PVS1_strong, PM2_supp | VUS |
| c.423-9A>G | ClinVar 469955 | Pathogenic | PP3, PM2_supp, PS3_mod | VUS | observed in 4 individuals worth 2 phenotype points | PP3, PM2_supp, PS3_mod, PS4_mod | Likely pathogenic |
| c.531+5_531+8del | ClinVar 537529 | Likely pathogenic | PP3, PM2_supp, PS3_mod, PS4_supp | VUS | observed in 5 individuals worth 3.5 phenotype points (56, 57) | PP3, PM2_supp, PS3_mod, PS4_mod | Likely pathogenic |
| c.531+5G>A | ClinVar 127305 | Pathogenic | PP3, PM2_supp, PS4_supp, PS1_mod | VUS | observed in 3 individuals worth 1.5 phenotype points; 2.5 phenotype points in total (58, 59) | PP3, PM2_supp, PS4_mod, PS1_mod | Likely pathogenic |
| c.531+6T>C | ClinVar 576816 | VUS | PP3, PM2_supp, PS3_mod | VUS | observed in 2 individuals worth 2 phenotype points | PP3, PM2_supp, PS3_mod, PS4_mod | Likely pathogenic |
| c.623A>G (p.Gln208Arg) | LOVD APC_000758 | Pathogenic | BP1, PM2_supp, PP1 | VUS | 1 phenotype point (60) | BP1, PM2_supp, PS4_supp, PP1 | VUS |
| c.645+2T>G | ClinVar 185659 | Likely pathogenic | PVS1_mod, PM2_supp | VUS | observed in 2 individuals worth 1 phenotype point | PVS1_mod, PM2_supp, PS4_supp | VUS |
| c.835-17A>G | ClinVar<br>822326 | Likely<br>pathogenic | PM2_supp, PS3_mod | VUS | observed in 3 individuals worth 0 phenotype points | PM2_supp, PS3_mod | VUS |
| c.835-7T>G | ClinVar<br>433614 | Likely<br>pathogenic | PP3, PM2_supp,<br>PS3_mod | VUS | observed in 1 individual worth 1 phenotype point | PP3, PM2_supp, PS3_mod,<br>PS4_supp | VUS |
| c.933G>C (p.Lys311Asn) | 1025291 | Likely<br>pathogenic | PVS1_supp,<br>PM2_supp | VUS | observed in 2 individuals worth 1.5 phenotype points (61) | PVS1_strong, PM2_supp,<br>PS4_supp | Likely<br>pathogenic |
| c.1042C>T (p.?) | ClinVar<br>955439 | Pathogenic | PVS1 | VUS | 1 phenotype point (62, 63); revised MAF criteria | PS4_supp, PM2_supp,<br>BS2_supp | VUS |
| c.1312+3A>C | ClinVar<br>486792 | Likely<br>pathogenic | PP3, PM2_supp,<br>PS1_mod | VUS | observed in 1 individual worth 1 phenotype point;<br>1.5 phenotype points in total (64) | PP3, PM2_supp, PS1_mod,<br>PS4_supp | VUS |
| c.1312+5G>C | ClinVar<br>265372 | Likely<br>pathogenic | PM2_supp,<br>PS4_supp, PS1_mod | VUS | observed in 2 individuals worth 2.5 phenotype points | PM2_supp, PS4_mod,<br>PS1_mod | VUS |
| c.1408+735A>T | LOVD<br>APC_001244 | Pathogenic | PM2_supp, PS3_mod,<br>PS4_supp | VUS | 1 phenotype point (65) | PS4_mod, PS3_mod,<br>PM2_supp | VUS |
| c.1409-5A>G | ClinVar<br>411406 | Pathogenic | PP3, PM2_supp,<br>PS3_mod, PS4_supp | VUS | observed in 5 individuals worth 3 phenotype points | PS3_mod, PS4_mod, PP3,<br>PM2_supp | Likely<br>pathogenic |
| c.1409-3T>G | ClinVar<br>485146 | Likely<br>pathogenic | PP3, PM2_supp,<br>PS3_mod | VUS | observed in 4 individuals worth 2 phenotype points (61) | PS3_mod, PS4_mod, PP3,<br>PM2_supp | Likely<br>pathogenic |
| c.1743G>C (p.Lys581Asn) | ClinVar<br>428153 | Likely<br>pathogenic | PVS1_strong,<br>PM2_supp | VUS | observed in 7 individuals worth 1 phenotype point | PVS1_strong, PS4_supp,<br>PM2_supp | Likely<br>pathogenic |
| c.1902T>G (p.Ser634Arg) | ClinVar<br>231954 | Likely<br>pathogenic | BP1, PS3_mod | VUS | observed in 2 individuals worth 0.5 phenotype points; revised MAF criteria and functional data | BP1, PM2_supp | VUS |
| c.3950A>G<br>(p.Glu1317Gly) | ClinVar<br>1319598 | VUS | BP1, PM2_supp | VUS | observed in 2 individuals worth 0 phenotype points | BP1, PM2_supp | VUS |
| c.4139C>T (p.Thr1380Ile) | ClinVar<br>233890 | VUS | BP1, PM2_supp,<br>PS4_supp | VUS | observed in 9 individuals worth 3 phenotype points | BP1, PM2_supp, PS4_mod | VUS |
| c.4735A>T (p.Ile1579Phe) | ClinVar<br>246402 | VUS | BP1, PM2_supp,<br>PS3_supp | VUS | observed in 4 individuals worth 1.5 phenotype points | BP1, PM2_supp, PS3_supp<br>PS4_supp | VUS |
| c.5038C>T (p.Gln1680Ter) | ClinVar<br>230520 | Likely<br>pathogenic | PVS1, BS1 | VUS | observed in 1 individual worth 0 phenotype points;<br>revised MAF criteria | PVS1, PM2_supp<sup>a</sup> | Likely<br>pathogenic |
| c.6905C>G<br>(p.Ser2302Ter) | ClinVar<br>428166 | Pathogenic | PVS1 | VUS | observed in 1 individual worth 0 phenotype points;<br>revised MAF criteria | PVS1, PM2_supp<sup>a</sup> | Likely<br>pathogenic |
| c.7489_7490insT<br>(p.Ser2497PhefsTer14) | ClinVar<br>653103 | Pathogenic | PVS1, BS1 | VUS | observed in 4 individuals worth 1 phenotype point;<br>revised MAF criteria | PVS1, PS4_supp,<br>PM2_supp<sup>a</sup> | Pathogenic |
| c.7798_7801del<br>(p.Gln2600ValfsTer15) | ClinVar<br>827255 | Pathogenic | PVS1, BS1 | VUS | observed in 2 individuals worth 0.5 phenotype points;<br>observed in 3 individuals worth 3 healthy individual points;<br>revised MAF criteria | PVS1, PM2_supp<sup>a</sup>,<br>BS2_supp | Likely<br>pathogenic |
| c.7803_7807del<br>(p.Ser2601ArgfsTer17) | ClinVar<br>648862 | Likely<br>pathogenic | PVS1 | VUS | observed in 1 individual worth 0 phenotype points;<br>revised MAF criteria | PVS1, PM2_supp<sup>a</sup> | Likely<br>pathogenic |

<sup>a</sup> Variants for which the reassessment of Minor allele frequency (MAF) criteria were relevant for the final evaluation as LP/P

## RESULTS

### Variant assessment using the reclassification algorithm based on VCEP *APC-*specific criteria

A total of 10,228 *APC* variants were analysed in this study, which included 190 (2%) structural and 10,038 (98%) non-structural variants (Supplementary Table 2). A total of 9,121 and 1,887 *APC* variants were present in ClinVar and LOVD, respectively (Figure 3); 780 (41% of LOVD) variants are shared between ClinVar and LOVD. The largest group in the ClinVar dataset is the VUS class (67%), followed by the B/LB group (20%) and P/LP group (14%). Most variants in LOVD are P/LP (76%), whereas VUS only account for 16% and B/LB variants for 8%.

**Figure 3.**
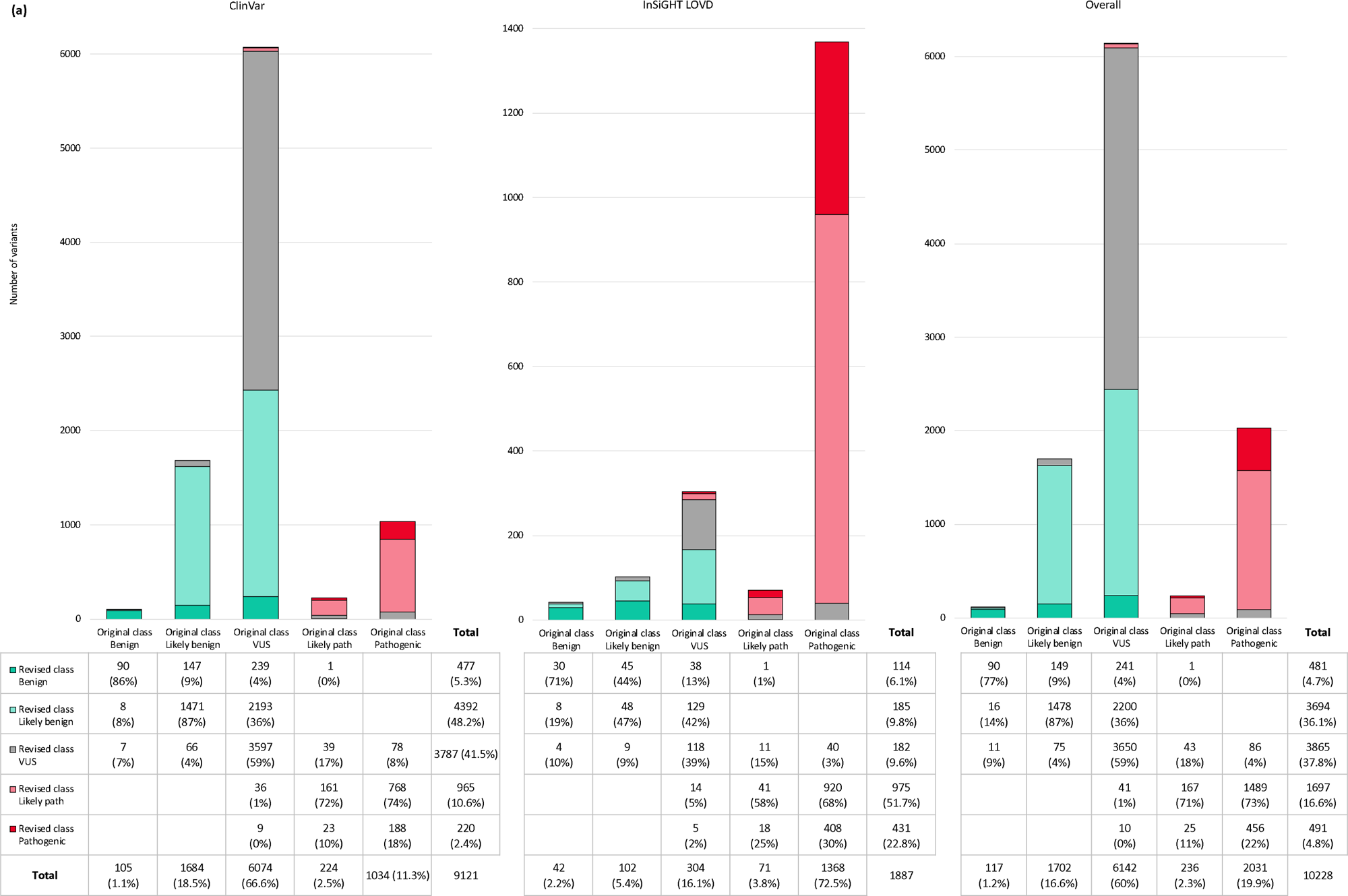
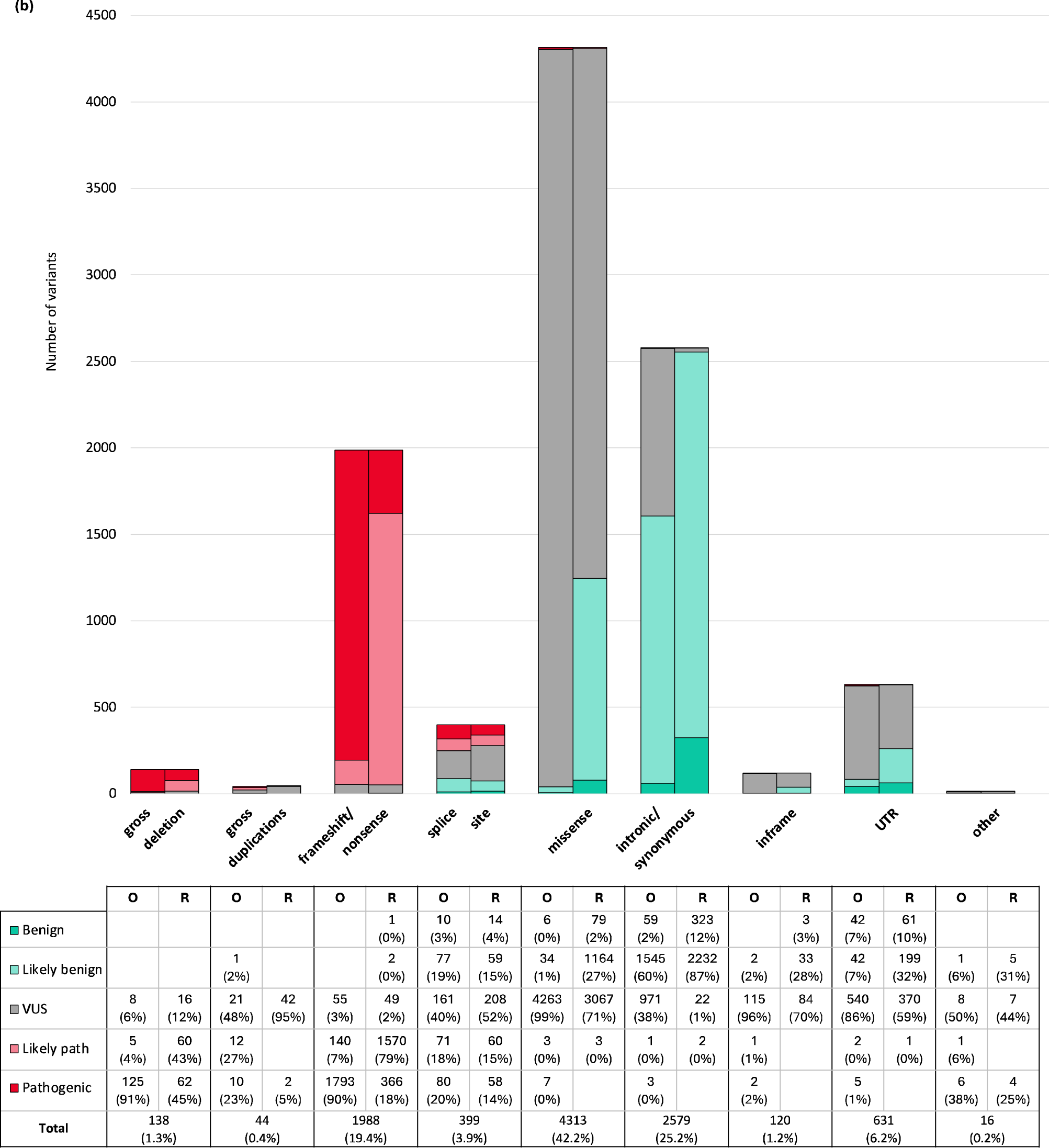
(a) Revised classification of all *APC* variants using the *APC-*specific criteria. Each column shows the number of variants with their original assertions in ClinVar, InSiGHT LOVD and the overall dataset. The coloured segments of each column represent the revised classification using the *APC*-specific criteria. **(b) Classification of all *APC* variants in the original database (O) and their revised classification (R) by variant type** Variants are broadly categorised into nine categories: 138 gross deletions, 44 gross duplications, 1988 frameshift/nonsense, 399 splice site, 4313 missense, 2579 intronic/synonymous, 120 in-frame, 631 UTR, and 16 other variants, which included start-loss, stop-loss, stop-retained, Alu and SVA retrotransposon insertions, inversions and complex variant.

Comparison between the original ClinVar or LOVD assertions and the revised classification are shown in Figure 3a. By applying the *APC-*specific criteria, in the ClinVar dataset, 4,869 variants were classified as B/LB (53%), 1,185 as P/LP (13%), and 3,787 as VUS (42%). In the LOVD dataset, 299 variants were classified as B/LB (16%), 1,406 as P/LP (75%), and 182 as VUS (10%). Notably, 94% of ClinVar variants with an initial classification of B/LB or P/LP remained in their respective benign and pathogenic categories after reclassification. Similarly, 96% LOVD variants were reclassified without significant clinical conflict. A considerable portion of previous P variants were downgraded to LP, which included 768 out of 1034 (74%) and 920 out of 1,368 (67%) previous P variants in ClinVar and LOVD, respectively. Remarkably, 40% of ClinVar VUS were reclassified as B/LB and 1% as P/LP, while in LOVD 55% VUS were reclassified as B/LB and 6% as P/LP. The percentage of VUS was reduced from 67% to 42% in ClinVar and from 16% to 10% in LOVD. As a whole, the total number of VUS in the combined dataset was reduced by 37% from 6,142 to 3,865 [38% in ClinVar (from 6,074 to 3,787) and 40% in LOVD (from 304 to 182)].

Only one previously LP variant was reclassified as LB by the *APC-*specific criteria: c.70C>T (p.Arg24Ter). Since this variant is 5’ of codon 49, PVS1 is not applicable. This variant was present in 6 out of 268,098 alleles with a MAF of 0.00002 in the gnomAD v.2.1.1 non-cancer population, meeting the initial BS1 specification (version 1.0.0). Meanwhile in the most recent version of the criteria (version 2.1.0), the *APC* VCEP has recommended the use of the “filtering allele frequency” (FAF) for BA1 and BS1, this value is not available for this variant (BS1 not met). Based on a survey among the VCEP members and a literature search (36, 37), 67 heterozygote carriers of c.70C>T were identified, which are reported either without or with a very mild polyposis phenotype (only 3 carriers fulfilled 0.5 phenotype points each; PS4_supporting). There are also more than 10 unaffected healthy individuals ≥ 50 years in this survey, fulfilling BS2. As a result, the variant was classified as LB and PS4_supporting was not considered as conflicting.

A number of previously B/LB or P/LP variants were reclassified as VUS, which included 73 B/LB (0.7%) and 117 P/LP (1.1%) variants in ClinVar, and 13 B/LB (0.1%) and 51 P/LP (0.05%) variants in LOVD, respectively. The reasons underlying these changes are outlined in Supplementary Table 3 and explored in further detail in the discussion section.

The distribution of variants by type is shown in Figure 3b with the majority being missense variants (42%), followed by synonymous or intronic variants (25%), and truncating (frameshift/nonsense) variants (19%). The original (O) and revised (R) pathogenicity class based on the algorithm was compared for each variant type. 97% of frameshift/nonsense variants are P/LP and 99% of synonymous and intronic variants are B/LB according to the *APC*-specific criteria. In the original ClinVar and LOVD assertions, 99% of putative missense variants are VUS. This proportion was reduced significantly to 71% using the *APC-*specific criteria, where approximately 30% of all missense variants were reclassified as B/LB. Application of the *APC*-specific criteria reduced the percentage of synonymous/intronic (at or beyond +7/-21 intronic positions) VUS from 38% to 1%, classifying the vast majority as B/LB. For variants flanking splice sites (within +7/-21 intronic positions), the range of classifications was similar before and after reclassification. 31% in-frame variants and 42% UTR variants were also classified as B/LB, while prior to reclassification they were mostly VUS. After application of the specific criteria, 3,865 variants remained VUS, which included 3,067 missense variants (79%), and low numbers of other variant types (Figure 3).

### Impact and usage of the *APC*-specific codes

The frequency of use of each *APC*-specific code is noted in Supplementary Table 4. All criteria were used at least once during the reclassification process. The most frequently applied codes were the two pathogenic criteria PM2_supporting (used for 69% of all 10,228 variants) and PVS1 (21%), and the four benign criteria BP1 (41%), BP4 (27%), BP7 (22%), and BS1 (20%), while on the other hand, half of the codes are used for less than 1% of variants. 2,192 of all 2568 truncating variants (85%) were assigned PVS1_variable (including frameshift, nonsense, spliceogenic, gross deletions/duplications and last nucleotide of exon G to non-G variants). In contrast, PS4_variable could only be applied in 455 of the 2,208 individuals (21%) with annotated phenotype description in LOVD. 3,145 variants (31% of all variants) were present in the gnomAD v.2.1.1 non-cancer population and/or the UK Biobank non-CRC dataset, of which 427 (14%; 4% of all variants) could be classified as benign just by the BA1 code alone and 135 (4%; 1% of all variants) as likely benign just by the BS1 code alone.

The most common criteria combination was BP1 and PM2_supporting: 2,622 variants (26%) were missense variants that were absent from population databases and had a consistent benign splice prediction, resulting in a VUS classification. The second most common criteria combination was PVS1 and PM2_supporting, which applied to 2,165 variants: 77% (1,667/2,165) of these variants (16% of all variants) were classified as LP solely based on the two codes. This was followed by BP4 and BP7 leading to a LB classification for 1,708 synonymous and intronic variants (17% of all variants). The combination of BS1 and BP1 was given to 1,064 missense variants (10% of all variants) which resulted in a LB classification. 155 variants (1.5% of all variants) were present in the non-CRC control dataset of UK Biobank, which fulfilled the definition for a healthy unaffected individual in the APC-specific criteria and allowed the assessment for BS2_variable.

### Further data mining and criteria review for a prioritised list of variants

After the initial application of the *APC*-specific criteria, a considerable fraction of VUS (63%) remained as expected. We selected 36 promising variants from these remaining VUS with some evidence for pathogenicity (details see Materials & Methods), which formed a prioritised list of variants for further review as outlined in the workflow (Figure 2).

In 11 truncating variants (Table 1, encompassing both Group 1 and 2), the relegation of their prior LP/P classification were due to their presence at very low frequencies in reference population databases, in this case the occurrence of one allele in a gnomAD non-cancer subpopulation. Depending on the denominator (i.e., size of the subpopulation), 7 previously LP/P truncating variants were precluded from the use of PM2_supporting and 4 even fulfilled threshold for BS1 using version 1.0.0 of the *APC*-specific criteria. To resolve this issue, the *APC* VCEP added a caveat to PM2_supporting in the criteria version 2.1.0 where the allele frequency threshold of ≤ 0.0003% (0.000003) is only used if the allele count is > 1. To tolerate singleton allele occurrence in gnomAD, the *APC* VCEP set an allele frequency of < 0.001% (0.00001) (lower than BS1) if the allele count is ≤ 1. Moreover, the *APC* VCEP recommended in criteria version 2.1.0 the use of the FAF for BA1 and BS1 to avoid the issue of singleton alleles satisfying the allele frequency criteria. This allowed the use of PM2_supporting and the reclassification of these 11 variants as LP/P as shown in Table 1.

For five variants that were originally P/LP in ClinVar or LOVD but reclassified as VUS by the algorithm, a more extensive data mining and literature review led to the return of P as their final classification (Group 1 in Table 1 and Figure 2). The previous pathogenic classification would indicate the observation of these variants in affected individuals on multiple occasions and a targeted search was finally informative.

For the remaining 31 variants (Group 2 in Table 1 and Figure 2), internal lab contributors provided available phenotypic information that allowed the upgrade of classification from VUS to P/LP for 11 (35%) variants (Group 2 in Table 1 and Figure 2). Five variants of Group 2 were evaluated as LP based on the reassessment of the MAF criteria, but no relevant phenotypic information was available. Overall, further data mining of selected representative variants resulted in the enhanced classification of 21 out of 36 variants (58%) into meaningful pathogenicity classes.

## DISCUSSION

The rising number of VUS in clinically actionable genes such as *APC* represents an important issue in the post-genomic era that hinders the translation of genetic diagnostics into improved health outcomes. In this study, we first identified the current landscape of *APC* variants with respect to their original assertions in the two most important international databases for *APC* variants: ClinVar and the InSiGHT LOVD. A striking difference was noted in the distribution of pathogenicity classes: while around two-thirds of ClinVar variants were originally VUS and 20% B/LB, roughly 70% of *APC* variants on LOVD were P/LP (Figure 3). This is not unexpected since variants submitted to LOVD are usually detected in patients with the relevant phenotype (i.e., clinically evident colorectal adenomatous polyposis) where the detection of PV is more likely and the primary focus of curators of locus-specific databases. Conversely, data in ClinVar is derived from the increased application of high-throughput sequencing methods in patients with less specific or unrelated phenotypes and healthy individuals has generated an extensive catalogue of rare variants in actionable genes, the majority of which is expected to be benign or have a low penetrance. However, in the absence of overwhelming evidence, they are usually conservatively classified as VUS.

This study was the first application of the full set of ClinGen-approved gene-specific ACMG/AMP guidelines (24) to a large set of variants identified worldwide. We set out to reclassify as many *APC* variants as possible with the aim to improve consistency and accuracy in *APC* variant classification. By consolidating *APC* variants in ClinVar and LOVD, we provide a holistic overview of most APC variants identified to date using gene-specific criteria in individuals with a spectrum of diseases. Other smaller scale studies used gene-/disease-specifications that covered only partial evidence domains (38), or meta-classification methods such as the multifactorial likelihood analysis (39, 40).

One of the major findings of this study is that the application of the *APC*-specific criteria reduced the number of VUS by 37% collectively in ClinVar and LOVD such that 2,277 variants could be revised into clinically meaningful classes. The majority (40%) of the former VUS were reclassified as B/LB (n= 2441) owing to their presence in reference population databases fulfilling MAF criteria (BA1/BS1) and/or computational data predicting no impact on protein function (BP1/BP4/BP7). As a result, these B/LB variants no longer need to be considered in the diagnosis of FAP and reported in genetic testing, which in turn minimises the anxiety and potential overtreatment in affected individuals worldwide (41). On the other hand, 51 previous VUS were reclassified as P/LP (0.8%), a result which confirms the genetic diagnosis of FAP and enables the timely management and predictive testing of all at-risk relatives. On the other hand, 95% and 94% of the previously B/LB and P/LP variants remained in the B/LB and P/LP classifications, respectively. These findings demonstrate that the application of the *APC*-specific criteria is highly effective in substantially improving the reclassification of VUS into clinically relevant pathogenicity classes while preserving the original interpretation of variants with existing evidence-based classifications.

The only variant [c.70C>T;(p.Arg24Ter)] with a clinically significant change from LP to LB was due to the indiscriminate use of PVS1 by the original submitters. Historically, variants are generally assumed to be pathogenic if they are protein truncating, without considering the impact of alternative start codon, alternative splicing, the preservation of relevant functional domains, and nonsense mediated decay. The *APC* VCEP conservatively defined that PVS1 is only applicable for truncating variants between codon 49 and 2645 inclusive (24). The *APC* variant c.70C>T;(p.Arg24Ter) is 5’ of that cut-off and has sufficient clinical evidence to suggest a LB classification as discussed in the results.

In the combined databases, a total of 128 P/LP variants (92 non-structural, 37 structural) and a total of 87 B/LB variants (86 structural, 1 non-structural) were reclassified as VUS (216 in total, 5% of previous non-VUS), which is clinically significant for the previous P/LP variants and will affect the diagnosis, counselling, and the predictive power of genetic testing (Supplementary Table 3). Among the 128 P/LP-to-VUS variants, 14 were gross deletions, which affected only the promoter region and/or exon 2 of the NM_000038.6 transcript (i.e., the first coding exon 1). The exact molecular consequences are difficult to ascertain, and they could not be properly classified in the absence of convincing clinical and segregation data although VCEP members thought they were likely pathogenic. Another 21 variants were gross duplications which had unknown or only presumed impact on the reading frame and lacking clinical data. While many *APC* gross duplications are indeed located in a tandem position (4, 42, 43), their impact on the transcript is unknown per se, especially if they extended beyond the open reading frame of the gene. Seven 5’UTR variants, nine truncating variants at the 5’ end and 7 truncating variants at the 3’ end of the gene were excluded from the application of PVS1 and were classified as VUS.

In addition to varying PVS1 application for frameshift and nonsense variants, we also noted that canonical ± 1/2 splice site changes and intronic variants flanking the splice sites (+7/-21 bp) were predominantly assessed as pathogenic by the submitters to LOVD and ClinVar, although splice predictions and transcript analyses might have suggested otherwise (i.e., weak native site, leaky splicing control etc). Among the 62 intronic variants in the vicinity of splice sites that were reclassified as VUS, 43 (69%) were originally LP/P and 19 (30%) were originally LB/B. The remaining variants included 55 deep intronic, synonymous, in-frame and UTR variants and 25 missense variants, where the reclassification as VUS was the result of a combination of scarcity of clinical data and non-contributory *in silico* predictions. These variants were better evaluated by the cautionary, conservative assessment intrinsic to the *APC*-specific criteria and highlights the importance of curating evidence from different domains.

In the *APC*-specific criteria, a range of evidence weight adjustments is specified as means of improve precision and quality. The lack of detailed evidence description in many studies meant that certain *APC*-specific criteria can only be applied at lower weights than what the submitters of ClinVar and LOVD might have intended. In this study, clinical data was only extracted from individuals with phenotype data in LOVD and for a selected number of variants where clinical data was essential for their reclassification into a non-VUS category. ClinVar and LOVD submitters may have additional variant information not apparent to this data-mining process. This is confirmed in the further curation of selected variants, where 56% of these VUS could again be reclassified into P/LP by additional data mining (Table 1). A key step was the request of further clinical information from our internal laboratory contributors, which substantially improved the interpretation of variants on the verge of being classified as LP/P.

A considerable number of previous P variants (75% in ClinVar and 68% in LOVD) were downgraded to LP (e.g. truncating variants fulfilling only PVS1 and PM2_supporting). This phenomenon is also known from other genes where gene-specific rules are being applied. Submitters may be overrating the quality of available pathogenic evidence, have additional evidence, applying the original moderate strength for PM2 or weighing the clinical information differently based on internal calibrations. A large-scale reclassification project is not able to screen for all available evidence for thousands of variants due to limited time and personal resources. Consequently, it cannot be excluded that some reclassified LP variants are in fact P, which has a great potential for future upgrade through diligent reporting of clinical information and data-sharing. In practical terms a LP classification has a posterior probability of pathogenicity of 0.9 to 0.99, which nonetheless demands clinical action when detected (44, 45).

After reclassification, 80% of the remaining VUS were presumed missense variants. The fundamental mechanism of *APC* pathogenicity is based on the loss of a large 3’ part of the protein which includes the relevant functional domains. In addition to further evidence of functional redundancy in the APC protein, the central and C-terminal domains of the APC protein are natively unfolded, all of which likely explains the resistance of the APC protein to missense variation (46) and the absolute predominance of truncating alterations among *APC* PV. The vast majority of the remaining missense *APC* VUS in ClinVar are likely benign incidental findings from non-targeted testing in individuals with unrelated phenotypes, for which *in silico* prediction tools and (massively parallel) functional assays are unlikely to contribute significantly to improve classifications. The recruitment of variant data from large reference population projects such as the UK Biobank on the other hand, is the key to determine the pathogenicity of these missense variants.

This reclassification endeavour was also considered a proof-of-concept study for the ongoing method of operation of VCEPs (Figure 2). The review and discussion of every single submitted constitutional variant in the *APC* gene is unrealistic with respect to the resources currently provided to a VCEP, until appropriate bioinformatic tools become available in the future. In this study, we developed a stratified variant curation process, whereby the relatively straightforward variants could be processed in batch and become candidates for fast-track VCEP review (e.g. variants fulfilling BA1 or BS1 plus BP1 without any conflicting classifications submitted to ClinVar or LOVD) in an updated variant curation interface (VCI) to streamline the variant approval process. We identified several prioritised groups of variants which can be the subject of further targeted literature review (Suppl Table 3), data-mining, and clinical data request from database submitters and internal lab contributors, a process to enhance variant interpretation as demonstrated in this study. These challenging variants can also form the basis for scientific follow-up studies to evaluate the causal relationships using additional investigations such as segregation or transcript analyses.

To our knowledge, this is the first study to undertake a large-scale approach to the interpretation of VUS in ClinVar and LOVD using ClinGen-approved gene-specific criteria. We developed and applied an algorithm for variant reclassification and demonstrated that the application of *APC*-specific criteria can substantially alleviate the burden of VUS, thereby laying the groundwork for a prospective streamlined expert panel approval of clinically actionable *APC* variants in the VCI. By using the VCEP specifications, diagnostic laboratories could reduce their rates of reporting VUS.

This study highlights the utility of a systematic, data-driven analysis using gene*-*specific ACMG/AMP criteria, complemented by further targeted data-mining and clinical data requests. By this approach, this study marks the initiation of a dynamic, long-term curation process for the *APC* VCEP. The suggested workflow also serves as a generalisable model of operation for other gene-/disease-specific variant interpretation initiatives, achieving accurate and highly efficient variant interpretation based on an array of carefully curated evidence. To further improve VUS interpretation and provide clinically informative variant classification beyond this approach, the availability of more population-based datasets, and user-friendly modes of sharing clinical and molecular data are needed; a challenge that has to be handled by the respective expert communities and data submitters.

## Data Availability

All data produced in the present work are contained in the manuscript

## Acknowledgements

This work was supported (not financially) by the European Reference Network on Genetic Tumour Risk Syndromes (ERN GENTURIS)-Project ID No 739547. ERN GENTURIS is partly co-funded by the European Union within the framework of the Third Health Programme “ERN-2016—Framework Partnership Agreement 2017-2021.”

