## Supplemental Table 1 for "Systematic large-scale application of ClinGen InSiGHT *APC*-specific ACMG/AMP variant classification criteria substantially alleviates the burden of variants of uncertain significance in ClinVar and LOVD databases"

**Supplementary Table 1 Status of ClinVar and all *APC* variant databases**

| Database name & URL | Curation & submission status | Date of Access | Submissions | Unique variant | Status |
| --- | --- | --- | --- | --- | --- |
| ClinVar [https://www.ncbi.nlm.nih.gov/clinvar/?term=APC[gene]](https://www.ncbi.nlm.nih.gov/clinvar/?term=APC%5bgene%5d) | Not curated, accepting submissions | 21 March 2022 | 18029 | 9121 | Active |
| Global Variome shared LOVD <https://www.lovd.nl/apc>; <https://databases.lovd.nl/shared/genes/APC> | Curated, accepting submissions | 12 May 2022 | 5663 | 1877 | Active |
| InSiGHT APC LOVD  <http://www.insight-database.org/genes/APC> | Merged with Global Variome shared LOVD | 12 May 2022 | 5663 | 1877 | Active |
| The UMD APC mutations database <http://www.umd.be/APC/> | Curated, not accepting submissions | 21 July 2022 | 3717 | 720 | Active |
| Argentina National Institute of Cancer  <http://www.inc.gob.ar/sither/genes/APC> | Curated, accepting submissions | 21 July 2022 | 161 | 48 | Active |
| Brazilian initiative on Precision Medicine  <http://bipmed.iqm.unicamp.br/genes/APC>  <http://bipmed.iqm.unicamp.br/snparray/genes/APC>  <http://bipmed.iqm.unicamp.br/snparray_hg19/genes/APC>  <http://bipmed.iqm.unicamp.br/snparray_296/genes/APC>  <http://bipmed.iqm.unicamp.br/wes_hg19/genes/APC> | Not curated, not accepting submissions | 21 July 2022 | 54  23  23  23  74 | 54  23  23  23  74 | Active |
| The APC mutation database <http://fap.taenzer.me/> | Inactive, URL not found |  |  |  | Inactive |
| Canadian Open Genetics Repository  [http://opengenetics.ca/#/brca/gene/APC](http://opengenetics.ca/) | Curated, not accepting submissions | 21 July 2022 |  | 259 | Active |
| CanVas – A Greek Cancer Patient Genetic Variation Resource  <http://ithaka.rrp.demokritos.gr/CanVaS/genes/APC> | Curated, accepting submissions | 21 July 2022 | 593 | 177 | Active |
| The Cyprus APC LOVD <http://db.cshg.org.cy/genes/APC> | Inactive, URL not found |  |  |  | Inactive |
| Dian Diagnostics & Zhejiang University Center for Genetic and Genomic Medicine <http://www.genomed.org/lovd2/home.php?select_db=APC> | Installation lost |  |  |  | Inactive |
| Iran Variation Database <http://genet.ir/variome/genes/APC> | Contained hidden *APC* variants only; LOVD installation partially broken |  |  |  | Inactive |
| Malaysian Node of the Human Variome Project Database  <http://www.kk.usm.my/LOVDv.3.0/genes/APC> | Curated, accepting submissions | 21 July 2022 | 35 | 29 | Active |
| MexVar <https://bipmed.fcm.unicamp.br/mexvar/genes/APC> | Inactive, URL not found |  |  |  | Inactive |
| Nicaragua APC <http://databases.lovd.ni/shared/genes/APC> | Inactive, URL not found |  |  |  | Inactive |
| Spain MDB <https://lovd3.isciii.es/genes/APC> | Did not contain any *APC* variant record |  |  |  | Inactive |
| Zhejiang University-Adinovo Center APC Database <http://databases.lovd.nl/genomed/home.php?select_db=APC> | Inactive, URL not found |  |  |  | Inactive |
| Other LOVD installation – LOVD3 whole genome datasets  <http://databases.lovd.nl/whole_genome/> | Not curated; not accepting submissions; imported from Exome Variant Server | 21 July 2022 | 321 | 321 | Active |
| Other LOVD installation – by the University of Melbourne <http://proteomics.bio21.unimelb.edu.au/lovd/genes/APC> | Not curated; inactive, not accepting submissions |  |  |  | Inactive |
