## Supplemental Table 3 for "Systematic large-scale application of ClinGen InSiGHT *APC*-specific ACMG/AMP variant classification criteria substantially alleviates the burden of variants of uncertain significance in ClinVar and LOVD databases"

**Supplementary Table 3 A list of all variants reclassified as VUS from clinically relevant classifications (B/LB/P/LP)**

*Caveat: Classification based on the ClinGen InSiGHT Hereditary Colorectal Cancer/Polyposis Expert Panel Specifications to the ACMG/AMP Variant Interpretation Guidelines for APC Version 1.0.0. Criteria applied in a stepwise fashion using a classification algorithm, not all available evidence may have been considered to arrive at current classification. All classifications are preliminary.*

| *Deletion at the extremities of the gene with unclear molecular consequences on the protein structure* | | | | | | | | | | | | | | | | | | | |
| --- | --- | --- | --- | --- | --- | --- | --- | --- | --- | --- | --- | --- | --- | --- | --- | --- | --- | --- | --- |
|  | **Database_ID** | **HGVSc; HGVSp** | **Predicted consequence** | | | | | | | | | **Prior classification** | | | | | **Reclassification by APC-specific criteria** | | |
|  | ClinVar 495348 | NM_000038.5:c.(?_-37541)_(-27791_?)del | deletion: promoter 1B | | | | | | | | | **Pathogenic** | | | | | VUS: PM2_supporting | | |
|  | ClinVar 832064 | NC_000005.10:g.(?_112707312)_(112755035_?)del | deletion: promoter 1B and 1A, exons 2 | | | | | | | | | **Pathogenic** | | | | | VUS: PM2_supporting | | |
|  | LOVD APC_001246 | NM_001127511.3:c.165+17816_166-18738delinsTGCTCTATGACCAATT | deletion: promoter 1B deletion | | | | | | | | | **Pathogenic** | | | | | VUS: PM2_supporting | | |
|  | ClinVar 580918 | NC_000005.9:g.112001178_112043328del42151 | deletion: promoter 1B partial | | | | | | | | | Likely pathogenic | | | | | VUS: PM2_supporting | | |
|  | ClinVar 433562 | NM_000038.5:c.-85-?_-19+?del | deletion: exon 1 | | | | | | | | | **Pathogenic** | | | | | VUS: PM2_supporting | | |
|  | LOVD APC_000526 | NM_000038.6:c.-85_(135+1_136-1) | deletion: exon 1-2 | | | | | | | | | **Pathogenic** | | | | | VUS: PS4_supporting, PM2_supporting | | |
|  | ClinVar 1049778 | NM_000038.6:c.-2_135+1824del | deletion: exon 2 | | | | | | | | | **Pathogenic** | | | | | VUS: PM2_supporting | | |
|  | ClinVar 1049324 | NM_000038.6:c.-2_135+1274del | deletion: exon 2 | | | | | | | | | **Pathogenic** | | | | | VUS: PM2_supporting | | |
|  | ClinVar 1049076 | NM_000038.6:c.-2_136-2903del | deletion: exon 2 | | | | | | | | | **Pathogenic** | | | | | VUS: PM2_supporting | | |
|  | ClinVar 584665 | NM_000038.6:c.1895_1958+28del | deletion: exon 15 partial | | | | | | | | | **Pathogenic** | | | | | VUS: PM2_supporting | | |
|  | ClinVar 433572 | NM_000038.5:c.3146-?_8532+?del | deletion: exon 16 partial | | | | | | | | | **Pathogenic** | | | | | VUS: PM2_supporting | | |
|  | ClinVar 433571 | NM_000038.5:c.2155-?_3960+?del | deletion: exon 16 partial | | | | | | | | | **Pathogenic** | | | | | VUS: PM2_supporting | | |
|  | ClinVar 1071358 | NC_000005.9:g.(?_112174702)_112203173del | deletion: exon 16 partial | | | | | | | | | **Pathogenic** | | | | | VUS: PM2_supporting | | |
|  | ClinVar 1071357 | NC_000005.9:g.(?_112173930)_112310702del | deletion: exon 16 partial | | | | | | | | | **Pathogenic** | | | | | VUS: PM2_supporting | | |
| *Large duplication and complex variants with unknown impact on the reading frame* | | | | | | | | | | | | | | | | | | | |
|  | **Database_ID** | **HGVSc; HGVSp** | | | **Predicted consequence** | | | | | | | | **Prior classification** | | | | | **Reclassification by APC-specific criteria** | |
|  | ClinVar 1067201 | NC_000005.9:g.(?_112072721)_(112090732_?)dup | | | duplication: promoter 1A, exon 2 | | | | | | | | **Likely pathogenic** | | | | | VUS: PM2_supporting | |
|  | ClinVar 584349 | NC_000005.9:g.(?_112072721)_(112090728_?)dup | | | duplication: promoter 1A, exon 2 | | | | | | | | **Likely pathogenic** | | | | | VUS: PM2_supporting | |
|  | ClinVar 469686 | NC_000005.9:g.(?_112072721)_(112111440_?)dup | | | duplication: promoter 1A, exons 2-5 | | | | | | | | **Likely pathogenic** | | | | | VUS: PM2_supporting | |
|  | ClinVar 1067204 | NC_000005.9:g.(?_112071797)_112137006dup | | | duplication: promoter 1A, exons 2-7, exon 8 partial | | | | | | | | **Likely pathogenic** | | | | | VUS: PM2_supporting | |
|  | ClinVar 1067082 | NC_000005.9:g.(?_112090582)_(112157694_?)dup | | | duplication: exon 1-11 | | | | | | | | **Likely pathogenic** | | | | | VUS: PM2_supporting | |
|  | ClinVar 58098 | GRCh38/hg38 5q15-22.3(chr5:96454445-114050905)x3 | | | duplication: exon 1-16 | | | | | | | | **Pathogenic** | | | | | VUS: PM2_supporting | |
|  | ClinVar 394550 | GRCh37/hg19 5q21.3-35.3(chr5:106716357-180687338)x3 | | | duplication: exon 1-16 | | | | | | | | **Pathogenic** | | | | | VUS: PM2_supporting | |
|  | ClinVar 425542 | GRCh37/hg19 5q15-35.3(chr5:94844077-178830410)x3 | | | duplication: exon 1-16 | | | | | | | | Likely benign | | | | | VUS: PM2_supporting | |
|  | ClinVar 688598 | GRCh37/hg19 5q14.3-23.3(chr5:89949118-129317455)x3 | | | duplication: exon 1-16 | | | | | | | | **Pathogenic** | | | | | VUS: PM2_supporting | |
|  | ClinVar 607687 | GRCh37/hg19 5p15.33-q35.3(chr5:25328-180693344)x3 | | | duplication: exon 1-16 | | | | | | | | **Pathogenic** | | | | | VUS: PM2_supporting | |
|  | ClinVar 607681 | GRCh37/hg19 5p15.33-q35.3(chr5:13648-180905029)x3 | | | duplication: exon 1-16 | | | | | | | | **Pathogenic** | | | | | VUS: PM2_supporting | |
|  | ClinVar 441919 | GRCh37/hg19 5p15.33-q35.3(chr5:113577-180719789)x3 | | | duplication: exon 1-16 | | | | | | | | **Pathogenic** | | | | | VUS: PM2_supporting | |
|  | ClinVar 441920 | GRCh37/hg19 5p15.33-q35.3(chr5:113577-180719789) | | | duplication: exon 1-16 | | | | | | | | **Pathogenic** | | | | | VUS: PM2_supporting | |
|  | ClinVar 417556 | NC_000005.9:g.(?_112090570)_(112157688_?)dup | | | duplication: exon 2-11 | | | | | | | | **Likely pathogenic** | | | | | VUS: PVS1_strong, PM2_supporting | |
|  | ClinVar 469688 | Single allele | | | duplication: exon 2-4 | | | | | | | | **Likely pathogenic** | | | | | VUS: PVS1_strong, PM2_supporting | |
|  | ClinVar 830510 | NC_000005.10:g.(?_112754891)_(112767400_?)dup | | | duplication: exons 2-4 | | | | | | | | **Likely pathogenic** | | | | | VUS: PVS1_strong, PM2_supporting | |
|  | ClinVar 1067200 | NC_000005.9:g.(?_112090582)_(112137086_?)dup | | | duplication: exon 2-8 | | | | | | | | **Likely pathogenic** | | | | | VUS: PVS1_strong, PM2_supporting | |
|  | LOVD APC_001753 | NM_000038.6:c.(135+1_136-1)_(422+1_423-1)dup | | | duplication: exon 3-4 | | | | | | | | **Pathogenic** | | | | | VUS: PVS1_strong, PM2_supporting | |
|  | LOVD APC_000750 | NM_000038.6:c.(422+1_423-1)_(531+1_532-1)dup | | | duplication: exon 4-5 | | | | | | | | **Pathogenic** | | | | | VUS: PVS1_strong, PM2_supporting | |
|  | LOVD APC_001782 | NM_000038.6:c.(834+1_835-1)_(1408+1_1409-1)dup | | | duplication: exon9-11 | | | | | | | | **Likely pathogenic** | | | | | VUS: PVS1_strong, PM2_supporting | |
|  | ClinVar 1067081 | NC_000005.9:g.(?_112170638)_(112170872_?)dup | | | duplication: exon 15 | | | | | | | | **Likely pathogenic** | | | | | VUS: PVS1_strong, PM2_supporting | |
|  | LOVD APC_001415 | NM_000038.6:c.1806_1817delinsN[300] | | | other | | | | | | | | **Pathogenic** | | | | | VUS: PM2_supporting | |
|  | LOVD APC_001953 | complex 3.9mb rearrangement | | | other | | | | | | | | **Likely pathogenic** | | | | | VUS: PS4_supporting, PM2_supporting | |
|  | ClinVar 243004 | NM_001127511.2:c.[-125delA;-195A>C] | | | other | | | | | | | | **Pathogenic** | | | | | VUS: PM2_supporting | |
| *Variants at the 5’ end of the gene and therefore excluded from the application of PVS1* | | | | | | | | | | | | | | | | | | | |
|  | **Database_ID** | **HGVSc; HGVSp** | | | | **Predicted consequences** | | | | **Prior classification** | | | | | **Reclassification by APC-specific criteria** | | | | |
|  | ClinVar 243007 | NM_001127511.3:c.-192A>T | | | | UTR | | | | **Pathogenic** | | | | | VUS: BP4, PM2_supporting | | | | |
|  | ClinVar 243006 | NM_001127511.3:c.-192A>G | | | | UTR | | | | **Likely pathogenic** | | | | | VUS: BS1 | | | | |
|  | ClinVar 652807 | NM_001127511.3:c.-192_-191delinsTAGCAAGGG | | | | UTR | | | | **Likely pathogenic** | | | | | VUS: PM2_supporting | | | | |
|  | ClinVar 243005 | NM_001127511.3:c.-191T>C | | | | UTR | | | | **Pathogenic** | | | | | VUS: BP4, PM2_supporting | | | | |
|  | LOVD APC_001802 | NM_000038.6:c.-190G>A | | | | UTR | | | | **pathogenic** | | | | | VUS: PM2_supporting | | | | |
|  | ClinVar 1050412 | NM_001127511.3:c.166-28469_166-27547del | | | | UTR | | | | **Pathogenic** | | | | | VUS: PM2_supporting | | | | |
|  | ClinVar 1050584 | NM_001127511.3:c.166-28467del | | | | UTR | | | | **Pathogenic** | | | | | VUS: BP4, PM2_supporting | | | | |
|  | ClinVar 537477 | NM_000038.6:c.14del; NP_000029.2:p.Ser5TyrfsTer6 | | | | frameshift | | | | **Pathogenic** | | | | | VUS: PM2_supporting | | | | |
|  | ClinVar 630969 | NM_000038.6:c.26_27insTTTA; NP_000029.2:p.Leu9PhefsTer7 | | | | frameshift | | | | **Pathogenic** | | | | | VUS: PM2_supporting | | | | |
|  | ClinVar 628229 | NM_000038.6:c.32_33insA; NP_000029.2:p.Gln12AlafsTer3 | | | | frameshift | | | | **Pathogenic** | | | | | VUS : PM2_supporting | | | | |
|  | ClinVar 970282 | NM_000038.6:c.55G>T; NP_000029.2:p.Glu19Ter | | | | nonsense | | | | **Pathogenic** | | | | | VUS: PM2_supporting | | | | |
|  | ClinVar 470090 | NM_000038.6:c.74_75del; NP_000029.2:p.Gln25ArgfsTer5 | | | | frameshift | | | | **Pathogenic** | | | | | VUS: PM2_supporting | | | | |
|  | ClinVar 428113 | NM_000038.6:c.93del; NP_000029.2:p.Asn32IlefsTer13 | | | | frameshift | | | | **Pathogenic** | | | | | VUS: PM2_supporting | | | | |
|  | ClinVar, LOVD 428154 | NM_000038.6:c.104del; NP_000029.2:p.Thr35LysfsTer10 | | | | frameshift | | | | **Pathogenic** | | | | | VUS: PM2_supporting | | | | |
|  | ClinVar 537502 | NM_000038.6:c.108del; NP_000029.2:p.Lys36AsnfsTer9 | | | | frameshift | | | | **Pathogenic** | | | | | VUS: PM2_supporting | | | | |
|  | ClinVar 579756 | NM_000038.6:c.132dup; NP_000029.2:p.Lys45GlufsTer5 | | | | frameshift | | | | **Pathogenic** | | | | | VUS: PM2_supporting | | | | |
| *Variants that satisfied PVS1, however were present at very low frequencies in population reference database (PM2_supporting not met)* | | | | | | | | | | | | | | | | | | | |
|  | **Database_ID** | **HGVSc; HGVSp** | | | | | | **Predicted consequences** | | | | | | | | **Prior classification** | | | **Reclassification by APC-specific criteria** |
|  | ClinVar 654864 | NM_000038.6:c.156del; NP_000029.2:p.Gly53GlufsTer17 | | | | | | frameshift | | | | | | | | **Pathogenic** | | | VUS: PVS1 |
|  | ClinVar, LOVD 934724 | NM_000038.6:c.203del; NP_000029.2:p.Leu68TyrfsTer2 | | | | | | frameshift | | | | | | | | **Pathogenic** | | | VUS: PVS1 |
|  | ClinVar, LOVD 411479 | NM_000038.6:c.471G>A; NP_000029.2:p.Trp157Ter | | | | | | nonsense | | | | | | | | **Pathogenic** | | | VUS: PVS1 |
|  | ClinVar, LOVD 955439 | NM_000038.6:c.1042C>T; NP_000029.2:p.Arg348Ter | | | | | | nonsense | | | | | | | | **Pathogenic** | | | VUS: PVS1 |
|  | ClinVar 438865 | NM_000038.6:c.1333C>T; NP_000029.2:p.Gln445Ter | | | | | | nonsense | | | | | | | | **Pathogenic** | | | VUS: PVS1, BS1 |
|  | ClinVar, LOVD 183857 | NM_000038.6:c.4669_4670del; NP_000029.2:p.Ile1557Ter | | | | | | frameshift | | | | | | | | **Pathogenic** | | | VUS: PVS1 |
|  | ClinVar, LOVD 230520 | NM_000038.6:c.5038C>T; NP_000029.2:p.Gln1680Ter | | | | | | nonsense | | | | | | | | **Likely pathogenic** | | | VUS: PVS1, BS1 |
|  | ClinVar 428166 | NM_000038.6:c.6905C>G; NP_000029.2:p.Ser2302Ter | | | | | | nonsense | | | | | | | | **Pathogenic** | | | VUS: PVS1 |
|  | ClinVar 653103 | NM_000038.6:c.7489_7490insT; NP_000029.2:p.Ser2497PhefsTer14 | | | | | | frameshift | | | | | | | | **Pathogenic** | | | VUS: PVS1, BS1 |
|  | ClinVar 827255 | NM_000038.6:c.7798_7801del; NP_000029.2:p.Gln2600ValfsTer15 | | | | | | frameshift | | | | | | | | **Pathogenic** | | | VUS: PVS1, BS1 |
|  | ClinVar 648862 | NM_000038.6:c.7803_7807del; NP_000029.2:p.Ser2601ArgfsTer17 | | | | | | frameshift | | | | | | | | **Likely pathogenic** | | | VUS: PVS1 |
| *Missense variants that are unable to be classified, mainly because minor allele frequency thresholds for BA1/BS1 are not met and lack of additional information* | | | | | | | | | | | | | | | | | | | |
|  | **Database ID** | **HGVSc; HGVSp** | | **Predicted consequences** | | | | | | **Prior classification** | | | | | **Reclassification by APC-specific criteria** | | | | |
|  | LOVD APC_001042 | NM_000038.6:c.446A>T; NP_000029.2:p.Asp149Val | | missense | | | | | | **pathogenic** | | | | | VUS: BP1, PM2_supporting, PS4_supporting | | | | |
|  | LOVD APC_000758 | NM_000038.6:c.623A>G; NP_000029.2:p.Gln208Arg | | missense | | | | | | **pathogenic** | | | | | VUS: BP1, PM2_supporting, PP1 | | | | |
|  | LOVD APC_000627 | NM_000038.6:c.1060C>T; NP_000029.2:p.Pro354Ser | | missense | | | | | | **pathogenic** | | | | | VUS: BP1, PM2_supporting | | | | |
|  | ClinVar 231954 | NM_000038.6:c.1902T>G; NP_000029.2:p.Ser634Arg | | missense | | | | | | **Likely pathogenic** | | | | | VUS: BP1, PS3_moderate | | | | |
|  | ClinVar 428167 | NM_000038.6:c.3077A>C; NP_000029.2:p.Asn1026Thr | | missense | | | | | | **Likely pathogenic** | | | | | VUS : PM2_supporting, PM5_supporting | | | | |
|  | ClinVar 802 | NM_000038.6:c.3359G>A; NP_000029.2:p.Gly1120Glu | | missense | | | | | | **Pathogenic** | | | | | VUS: BP1, PM2_supporting | | | | |
|  | ClinVar 817 | NM_000038.6:c.4183A>T; NP_000029.2:p.Ser1395Cys | | missense | | | | | | **Pathogenic** | | | | | VUS: BP1, PM2_supporting | | | | |
|  | LOVD APC_000208 | NM_000038.6:c.4549C>G; NP_000029.2:p.Gln1517Glu | | missense | | | | | | **pathogenic** | | | | | VUS: BP1, PM2_supporting, PS4_supporting | | | | |
|  | LOVD APC_001630 | NM_000038.6:c.6257C>A; NP_000029.2:p.Pro2086Gln | | missense | | | | | | **Likely pathogenic** | | | | | VUS: BP1, PM2_supporting | | | | |
|  | ClinVar 486771 | NM_000038.6:c.743A>G; NP_000029.2:p.Asn248Ser | | missense | | | | | | Likely benign | | | | | VUS: BP1 | | | | |
|  | ClinVar 827113 | NM_000038.6:c.754A>G; NP_000029.2:p.Thr252Ala | | missense | | | | | | Likely benign | | | | | VUS : BP1, PM2_supporting | | | | |
|  | ClinVar 487015 | NM_000038.6:c.2581G>A; NP_000029.2:p.Gly861Ser | | missense | | | | | | Likely benign | | | | | VUS: BP1, PM2_supporting | | | | |
|  | LOVD APC_001944 | NM_000038.6:c.2605A>G; NP_000029.2:p.Asn869Asp | | missense | | | | | | Likely benign | | | | | VUS: BP1, PM2_supporting | | | | |
|  | ClinVar 617980 | NM_000038.6:c.2651C>T; NP_000029.2:p.Ala884Val | | missense | | | | | | Likely benign | | | | | VUS: BP1, PM2_supporting | | | | |
|  | ClinVar 428175 | NM_000038.6:c.2909G>C; NP_000029.2:p.Ser970Thr | | missense | | | | | | Likely benign | | | | | VUS: BP1, PM2_supporting | | | | |
|  | LOVD APC_001667 | NM_000038.6:c.3290A>G; NP_000029.2:p.Glu1097Gly | | missense | | | | | | Likely benign | | | | | VUS: BP1, PM2_supporting | | | | |
|  | ClinVar 482305 | NM_000038.6:c.3608G>T; NP_000029.2:p.Gly1203Val | | missense | | | | | | Likely benign | | | | | VUS: BP1, PM2_supporting | | | | |
|  | LOVD APC_001945 | NM_000038.6:c.3930G>T; NP_000029.2:p.Lys1310Asn | | missense | | | | | | Likely benign | | | | | VUS: BP1, PM2_supporting | | | | |
|  | LOVD APC_001676 | NM_000038.6:c.4316C>G; NP_000029.2:p.Pro1439Arg | | missense | | | | | | Likely benign | | | | | VUS: BP1, PM2_supporting | | | | |
|  | ClinVar 824838 | NM_000038.6:c.4370C>T; NP_000029.2:p.Ala1457Val | | missense | | | | | | Likely benign | | | | | VUS: BP1, PM2_supporting | | | | |
|  | LOVD APC_001865 | NM_000038.6:c.5249T>C; NP_000029.2:p.Val1750Ala | | missense | | | | | | Likely benign | | | | | VUS: BP1, PM2_supporting | | | | |
|  | ClinVar 482255 | NM_000038.6:c.5651C>G; NP_000029.2:p.Ala1884Gly | | missense | | | | | | Likely benign | | | | | VUS: BP1 | | | | |
|  | LOVD APC_001947 | NM_000038.6:c.5731C>A; NP_000029.2:p.Gln1911Lys | | missense | | | | | | Likely benign | | | | | VUS: BP1, PM2_supporting | | | | |
|  | ClinVar 617981 | NM_000038.6:c.5839A>C; NP_000029.2:p.Thr1947Pro | | missense | | | | | | Likely benign | | | | | VUS: BP1, PM2_supporting | | | | |
|  | ClinVar 236654 | NM_000038.6:c.8255A>C; NP_000029.2:p.Asn2752Thr | | missense | | | | | | Benign | | | | | VUS: BP1 | | | | |
| *Variants in the flanking intronic region with unknown consequences* | | | | | | | | | | | | | | | | | | | |
|  | **Database ID** | **HGVSc; HGVSp** | | **Predicted consequences** | | | | | **Prior classification** | | | **Reclassification by APC-specific criteria** | | | | | | | |
|  | ClinVar 482476 | NM_000038.6:c.135+1G>T | | splice | | | | | **Likely pathogenic** | | | VUS: PP3, PM2_supporting | | | | | | | |
|  | ClinVar 490194 | NM_000038.6:c.135+2T>C | | splice | | | | | **Likely pathogenic** | | | VUS: PP3, PM2_supporting | | | | | | | |
|  | ClinVar, LOVD 469955 | NM_000038.6:c.423-9A>G | | splice | | | | | **Pathogenic** | | | VUS: PP3, PM2_supporting, PS3_moderate | | | | | | | |
|  | LOVD APC_000602 | NM_000038.6:c.423-6_424delinsGAAGCAAGATCAG | | splice | | | | | **pathogenic** | | | VUS: PM2_supporting, PS4_supporting | | | | | | | |
|  | LOVD APC_000624 | NM_000038.6:c.531+1del | | splice | | | | | **pathogenic** | | | VUS: PP3, PM2_supporting, PM6, PS4_supporting | | | | | | | |
|  | ClinVar, LOVD 537529 | NM_000038.6:c.531+5_531+8del | | splice | | | | | **Likely pathogenic** | | | VUS: PP3, PM2_supporting, PS3_moderate, PS4_supporting | | | | | | | |
|  | ClinVar, LOVD 428099 | NM_000038.6:c.531+3A>C | | splice | | | | | **Likely pathogenic** | | | VUS: PP3, PM2_supporting | | | | | | | |
|  | ClinVar 127305 | NM_000038.6:c.531+5G>A | | splice | | | | | **Pathogenic** | | | VUS: PP3, PM2_supporting, PS4_supporting, PS1_moderate | | | | | | | |
|  | LOVD APC_001369 | NM_000038.6:c.532-2_532-1insAAAC | | splice | | | | | **pathogenic** | | | VUS: BP4, PM2_supporting | | | | | | | |
|  | LOVD APC_000429 | NM_000038.6:c.645+1G>C | | splice | | | | | **pathogenic** | | | VUS: PVS1_moderate, PM2_supporting, PS1_moderate | | | | | | | |
|  | LOVD APC_001803 | NM_000038.6:c.645+2T>C | | splice | | | | | **pathogenic** | | | VUS: PVS1_moderate, PM2_supporting, PS1_moderate | | | | | | | |
|  | ClinVar 185659 | NM_000038.6:c.645+2T>G | | splice | | | | | **Likely pathogenic** | | | VUS: PVS1_moderate, PM2_supporting, PS1_moderate | | | | | | | |
|  | ClinVar 822326 | NM_000038.6:c.835-17A>G | | splice | | | | | **Likely pathogenic** | | | VUS: PM2_supporting, PS3_moderate | | | | | | | |
|  | ClinVar, LOVD 433614 | NM_000038.6:c.835-7T>G | | splice | | | | | **Likely pathogenic** | | | VUS: PP3, PM2_supporting, PS3_moderate | | | | | | | |
|  | LOVD APC_000327 | NM_000038.6:c.933+1del | | splice | | | | | **pathogenic** | | | VUS: PP3, PM2_supporting, PS4_supporting | | | | | | | |
|  | ClinVar 181775 | NM_000038.6:c.1312+3_1312+4del | | splice | | | | | **Pathogenic** | | | VUS: PP3, PM2_supporting | | | | | | | |
|  | ClinVar 486792 | NM_000038.6:c.1312+3A>C | | splice | | | | | **Likely pathogenic** | | | VUS: PP3, PM2_supporting, PS1_moderate | | | | | | | |
|  | LOVD APC_001939 | NM_000038.6:c.1312+4_1312+19del | | splice | | | | | **pathogenic** | | | VUS: BP4, PM2_supporting | | | | | | | |
|  | ClinVar, LOVD 265372 | NM_000038.6:c.1312+5G>C | | splice | | | | | **Likely pathogenic** | | | VUS: PM2_supporting, PS4_supporting, PS1_moderate | | | | | | | |
|  | LOVD APC_001275 | NM_000038.6:c.1312+8C>T | | splice | | | | | **pathogenic** | | | VUS: BP4, PM2_supporting | | | | | | | |
|  | ClinVar 1066995 | NM_000038.6:c.1313-2A>C | | splice | | | | | **Likely pathogenic** | | | VUS: PP3, PM2_supporting | | | | | | | |
|  | ClinVar 578480 | NM_000038.6:c.1313-2A>G | | splice | | | | | **Likely pathogenic** | | | VUS: PP3, PM2_supporting, PS3_moderate | | | | | | | |
|  | ClinVar 1339656 | NM_000038.6:c.1313-1G>T | | splice | | | | | **Likely pathogenic** | | | VUS: PP3, PM2_supporting | | | | | | | |
|  | ClinVar 1323307 | NM_000038.6:c.1313-1G>C | | splice | | | | | **Pathogenic** | | | VUS: PP3, PM2_supporting | | | | | | | |
|  | LOVD APC_000792 | NM_000038.6:c.1408+7C>G | | splice | | | | | **pathogenic** | | | VUS: BP4, PM2_supporting, PS4_supporting | | | | | | | |
|  | ClinVar, LOVD 411406 | NM_000038.6:c.1409-5A>G | | splice | | | | | **Pathogenic** | | | VUS: PP3, PM2_supporting, PS3_moderate, PS4_supporting | | | | | | | |
|  | ClinVar, LOVD 485146 | NM_000038.6:c.1409-3T>G | | splice | | | | | **Likely pathogenic** | | | VUS: PP3, PM2_supporting, PS3_moderate | | | | | | | |
|  | LOVD APC_001291 | NM_000038.6:c.1548_1548+1delinsTT | | splice | | | | | **pathogenic** | | | VUS: BP4, PM2_supporting | | | | | | | |
|  | LOVD APC_001083 | NM_000038.6:c.1548+1del | | splice | | | | | **pathogenic** | | | VUS: PP3, PM2_supporting | | | | | | | |
|  | LOVD APC_001292 | NM_000038.6:c.1548+1_1548+9del | | splice | | | | | **pathogenic** | | | VUS: PP3, PM2_supporting | | | | | | | |
|  | ClinVar 265375 | NM_000038.6:c.1548+3_1548+4del | | splice | | | | | **Likely pathogenic** | | | VUS: PP3, PM2_supporting | | | | | | | |
|  | LOVD APC_001727 | NM_000038.6:c.1549-3C>G | | splice | | | | | **Likely pathogenic** | | | VUS: PP3, PM2_supporting | | | | | | | |
|  | LOVD APC_000214 | NM_000038.6:c.1627-8A>G | | splice | | | | | **pathogenic** | | | VUS: PP3, PM2_supporting | | | | | | | |
|  | LOVD APC_000321 | NM_000038.6:c.1742A>G; NP_000029.2:p.Lys581Arg | | splice | | | | | **pathogenic** | | | VUS: PM2_supporting, PS3_moderate | | | | | | | |
|  | ClinVar 428153 | NM_000038.6:c.1743G>C; NP_000029.2:p.Lys581Asn | | splice | | | | | **Likely pathogenic** | | | VUS: PVS1_strong, PM2_supporting | | | | | | | |
|  | LOVD APC_001411 | NM_000038.6:c.1743+1del | | splice | | | | | **pathogenic** | | | VUS: PP3, PM2_supporting | | | | | | | |
|  | LOVD APC_000296 | NM_000038.6:c.1744-17_1744-5delinsTC | | splice | | | | | **pathogenic** | | | VUS: BP4, PM2_supporting | | | | | | | |
|  | ClinVar 664704 | NM_000038.6:c.1744-6_1744-4delinsAG | | splice | | | | | **Pathogenic** | | | VUS: PP3, PM2_supporting | | | | | | | |
|  | ClinVar 433625 | NM_000038.6:c.1744-4C>G | | splice | | | | | **Likely pathogenic** | | | VUS: PM2_supporting | | | | | | | |
|  | ClinVar 819988 | NM_000038.6:c.1744-3T>G | | splice | | | | | **Pathogenic** | | | VUS: PP3, PM2_supporting | | | | | | | |
|  | LOVD APC_001467 | NM_000038.6:c.1954_1958+15del | | splice | | | | | **pathogenic** | | | VUS: BP4, PM2_supporting | | | | | | | |
|  | LOVD APC_001445 | NM_000038.6:c.1958+1del | | splice | | | | | **pathogenic** | | | VUS: PP3, PM2_supporting | | | | | | | |
|  | ClinVar 265560 | NM_000038.6:c.1958+3A>T | | splice | | | | | **Likely pathogenic** | | | VUS: PP3, PM2_supporting, PS1_moderate | | | | | | | |
|  | ClinVar 439406 | NM_000038.6:c.-18-13T>G | | splice | | | | | Benign | | | VUS: PM2_supporting | | | | | | | |
|  | ClinVar 672308 | NM_000038.6:c.135+6A>G | | splice | | | | | Likely benign | | | VUS: BP4, PM2_supporting | | | | | | | |
|  | ClinVar 1130754 | NM_000038.6:c.135+7A>G | | splice | | | | | Likely benign | | | VUS: BP4, PM2_supporting | | | | | | | |
|  | ClinVar 918312 | NM_000038.6:c.136-13T>G | | splice | | | | | Likely benign | | | VUS: BP4, PM2_supporting | | | | | | | |
|  | ClinVar 490196 | NM_000038.6:c.136-12T>C | | splice | | | | | Likely benign | | | VUS: BP4, PM2_supporting | | | | | | | |
|  | ClinVar 490239 | NM_000038.6:c.221-11A>T | | splice | | | | | Likely benign | | | VUS: BP4, PM2_supporting | | | | | | | |
|  | LOVD APC_001759 | NM_000038.6:c.423-18_423-17insA | | splice | | | | | benign | | | VUS: BP4, PM2_supporting | | | | | | | |
|  | ClinVar 181780 | NM_000038.6:c.423-17_423-16insT | | splice | | | | | Benign | | | VUS: BP4, PM2_supporting | | | | | | | |
|  | ClinVar 1143398 | NM_000038.6:c.423-9A>T | | splice | | | | | Likely benign | | | VUS: BP4, PM2_supporting | | | | | | | |
|  | ClinVar 1116379 | NM_000038.6:c.423-8A>T | | splice | | | | | Likely benign | | | VUS: BP4, PM2_supporting | | | | | | | |
|  | ClinVar 1149314 | NM_000038.6:c.423-4A>T | | splice | | | | | Likely benign | | | VUS: BP4, PM2_supporting | | | | | | | |
|  | ClinVar 537617 | NM_000038.6:c.532-7G>T | | splice | | | | | Likely benign | | | VUS: BP4 | | | | | | | |
|  | ClinVar 380375 | NM_000038.6:c.532-7G>C | | splice | | | | | Likely benign | | | VUS: BP4 | | | | | | | |
|  | ClinVar 1110067 | NM_000038.6:c.645+7T>C | | splice | | | | | Likely benign | | | VUS: BP4, PM2_supporting | | | | | | | |
|  | ClinVar 1137468 | NM_000038.6:c.645+8A>C | | splice | | | | | Likely benign | | | VUS: BP4, PM2_supporting | | | | | | | |
|  | ClinVar 926761 | NM_000038.6:c.645+8A>T | | splice | | | | | Likely benign | | | VUS: BP4, PM2_supporting | | | | | | | |
|  | ClinVar 793315 | NM_000038.6:c.646-8T>C | | splice | | | | | Likely benign | | | VUS: BP4, PM2_supporting | | | | | | | |
|  | ClinVar 490358 | NM_000038.6:c.729+3T>A | | splice | | | | | Likely benign | | | VUS: BP4, PM2_supporting | | | | | | | |
|  | ClinVar 1103461 | NM_000038.6:c.729+8A>G | | splice | | | | | Likely benign | | | VUS: BP4, PM2_supporting | | | | | | | |
| *Deep intronic, synonymous, in-frame variants and variants in the UTR that are unable to be classified due to missing additional information* | | | | | | | | | | | | | | | | | | | |
|  | **Database_ID** | **HGVSc; HGVSp** | | | | | **Predicted consequences** | | | | **Prior classification** | | | **Reclassification by APC-specific criteria** | | | | | |
|  | LOVD APC_001244 | NM_000038.6:c.1408+735A>T | | | | | intron | | | | **pathogenic** | | | VUS: PM2_supporting, PS3_moderate, PS4_supporting | | | | | |
|  | ClinVar 823173 | NM_000038.6:c.933+829A>G | | | | | intron | | | | **Likely pathogenic** | | | VUS: PM2_supporting | | | | | |
|  | ClinVar 1111220 | NM_001127511.3:c.-199C>T | | | | | UTR | | | | Likely benign | | | VUS: BP4, PM2_supporting | | | | | |
|  | ClinVar 1169564 | NM_001127511.3:c.-167_-166insG | | | | | UTR | | | | Benign | | | VUS: BP4, PM2_supporting | | | | | |
|  | ClinVar 641353 | NM_001127511.3:c.-134_-133insGGG | | | | | UTR | | | | Likely benign | | | VUS: BP4, PM2_supporting | | | | | |
|  | ClinVar 1164593 | NM_001127511.3:c.-133_-132delinsGT | | | | | UTR | | | | Benign | | | VUS: PM2_supporting | | | | | |
|  | LOVD APC_001812 | NM_001127511.3:c.-126dup | | | | | UTR | | | | Likely benign | | | VUS: PM2_supporting | | | | | |
|  | ClinVar 1316326 | NM_001127511.3:c.-126_-125insGA | | | | | UTR | | | | Likely benign | | | VUS: BP4, PM2_supporting | | | | | |
|  | ClinVar 469825 | NM_001127511.3:c.-124C>G | | | | | UTR | | | | Benign | | | VUS: BP4, PM2_supporting | | | | | |
|  | LOVD APC_001815 | NM_001127511.3:c.15G>C; NP_001120983.2:p.Gly6= | | | | | UTR | | | | Likely benign | | | VUS: PM2_supporting | | | | | |
|  | ClinVar 918662 | NM_000038.6:c.730-19G>T | | | | | intron | | | | Likely benign | | | VUS: BP4, PM2_supporting | | | | | |
|  | ClinVar 796802 | NM_000038.6:c.834+7A>G | | | | | splice | | | | Likely benign | | | VUS: BP4, PM2_supporting | | | | | |
|  | ClinVar 628015 | NM_000038.6:c.835-20A>C | | | | | intron | | | | Likely benign | | | VUS: BP4, PM2_supporting | | | | | |
|  | ClinVar 381344 | NM_000038.6:c.835-15G>A | | | | | splice | | | | Likely benign | | | VUS: BP4, PM2_supporting | | | | | |
|  | ClinVar 1332031 | NM_000038.6:c.835-13G>A | | | | | splice | | | | Likely benign | | | VUS: BP4, PM2_supporting | | | | | |
|  | ClinVar 1115430 | NM_000038.6:c.835-10T>C | | | | | splice | | | | Likely benign | | | VUS: PM2_supporting | | | | | |
|  | LOVD APC_000439 | NM_000038.6:c.933G>A; NP_000029.2:p.Lys311= | | | | | splice | | | | benign | | | VUS: PVS1_supporting, PM2_supporting | | | | | |
|  | ClinVar 927827 | NM_000038.6:c.934-7C>T | | | | | splice | | | | Likely benign | | | VUS: BP4, PM2_supporting | | | | | |
|  | ClinVar 823565 | NM_000038.6:c.993G>T; NP_000029.2:p.Ser331= | | | | | synonymous | | | | Likely benign | | | VUS | | | | | |
|  | ClinVar 734263 | NM_000038.6:c.1071C>A; NP_000029.2:p.Ile357= | | | | | synonymous | | | | Likely benign | | | VUS: PM2_supporting | | | | | |
|  | ClinVar 1119186 | NM_000038.6:c.1312+8C>G | | | | | splice | | | | Likely benign | | | VUS: BP4, PM2_supporting | | | | | |
|  | ClinVar 928340 | NM_000038.6:c.1313-15G>T | | | | | splice | | | | Likely benign | | | VUS: BP4, PM2_supporting | | | | | |
|  | ClinVar 921987 | NM_000038.6:c.1313-14del | | | | | splice | | | | Likely benign | | | VUS: BP4, PM2_supporting | | | | | |
|  | ClinVar 918459 | NM_000038.6:c.1313-13T>C | | | | | splice | | | | Likely benign | | | VUS: BP4 | | | | | |
|  | ClinVar 416761 | NM_000038.6:c.1313-8T>A | | | | | splice | | | | Likely benign | | | VUS: BP4, PM2_supporting | | | | | |
|  | ClinVar 819130 | NM_000038.6:c.1407A>G; NP_000029.2:p.Leu469= | | | | | splice | | | | Likely benign | | | VUS: BP4 | | | | | |
|  | ClinVar 919829 | NM_000038.6:c.1408+7C>T | | | | | splice | | | | Likely benign | | | VUS: BP4, PM2_supporting | | | | | |
|  | ClinVar 1128391 | NM_000038.6:c.1408+8A>T | | | | | splice | | | | Likely benign | | | VUS: BP4, PM2_supporting | | | | | |
|  | ClinVar 388511 | NM_000038.6:c.1408+8A>G | | | | | splice | | | | Likely benign | | | VUS: BP4, PM2_supporting | | | | | |
|  | ClinVar 371848 | NM_000038.6:c.1409-17T>G | | | | | splice | | | | Likely benign | | | VUS: BP4 | | | | | |
|  | ClinVar 923782 | NM_000038.6:c.1409-16G>A | | | | | splice | | | | Likely benign | | | VUS: BP4 | | | | | |
|  | ClinVar 922204 | NM_000038.6:c.1409-16G>C | | | | | splice | | | | Likely benign | | | VUS: BP4, PM2_supporting | | | | | |
|  | ClinVar 508018 | NM_000038.6:c.1409-13C>G | | | | | splice | | | | Likely benign | | | VUS: BP4, PM2_supporting | | | | | |
|  | ClinVar 922965 | NM_000038.6:c.1410G>T; NP_000029.2:p.Gly470= | | | | | splice | | | | Likely benign | | | VUS: BP4, PM2_supporting | | | | | |
|  | ClinVar 630114 | NM_000038.6:c.1626+8T>G | | | | | splice | | | | Likely benign | | | VUS: BP4, PM2_supporting | | | | | |
|  | ClinVar 490219 | NM_000038.6:c.1626+8T>C | | | | | splice | | | | Likely benign | | | VUS: BP4, PM2_supporting | | | | | |
|  | ClinVar 490220 | NM_000038.6:c.1627-17A>G | | | | | splice | | | | Likely benign | | | VUS: PM2_supporting | | | | | |
|  | ClinVar 627826 | NM_000038.6:c.1627-16A>C | | | | | splice | | | | Likely benign | | | VUS: BP4, PM2_supporting | | | | | |
|  | ClinVar 490223 | NM_000038.6:c.1744-20C>T | | | | | intron | | | | Likely benign | | | VUS: BP4, PM2_supporting | | | | | |
|  | ClinVar 548881 | NM_000038.6:c.1744-14_1744-13del | | | | | splice | | | | Likely benign | | | VUS: BP4, PM2_supporting | | | | | |
|  | LOVD APC_001729 | NM_000038.6:c.1744-11T>G | | | | | splice | | | | Likely benign | | | VUS: PM2_supporting | | | | | |
|  | ClinVar 516255 | NM_000038.6:c.1744-10T>C | | | | | splice | | | | Likely benign | | | VUS: BP4, PM2_supporting | | | | | |
|  | LOVD APC_000607 | NM_000038.6:c.1869G>T; NP_000029.2:p.Arg623= | | | | | synonymous | | | | benign | | | VUS: BP4, BP7, PM2_supporting, PS3_moderate, PS4 | | | | | |
|  | ClinVar 627857 | NM_000038.6:c.1959-18C>G | | | | | intron | | | | Likely benign | | | VUS: BP4, PM2_supporting | | | | | |
|  | ClinVar 630908 | NM_000038.6:c.1959-17T>C | | | | | splice | | | | Likely benign | | | VUS: BP4, PM2_supporting | | | | | |
|  | LOVD APC_000458 | NM_000038.6:c.1959G>C; NP_000029.2:p.Arg653Ser | | | | | splice | | | | benign | | | VUS: PM2_supporting | | | | | |
|  | ClinVar 389718 | NM_000038.6:c.1962A>G; NP_000029.2:p.Gln654= | | | | | synonymous | | | | Likely benign | | | VUS: PM2_supporting | | | | | |
|  | ClinVar 733258 | NM_000038.6:c.2031C>G; NP_000029.2:p.Val677= | | | | | synonymous | | | | Likely benign | | | VUS | | | | | |
|  | ClinVar 918747 | NM_000038.6:c.4966_4967delinsAG; NP_000029.2:p.Ser1656= | | | | | synonymous | | | | Likely benign | | | VUS: PM2_supporting | | | | | |
|  | ClinVar 439411 | NM_000038.6:c.5265_5268delinsATCG; NP_000029.2:p.AlaSer1755= | | | | | synonymous | | | | Benign | | | VUS: PM2_supporting | | | | | |
|  | ClinVar 757607 | NM_000038.6:c.7404_7406del; NP_000029.2:p.Ser2469del | | | | | inframe | | | | Likely benign | | | VUS: BP4, PM2_supporting | | | | | |
|  | ClinVar 802150 | NM_000038.6:c.*415_*414insAAAAAA | | | | | UTR | | | | Likely benign | | | VUS: PM2_supporting | | | | | |
|  | ClinVar 217930 | NM_000038.6:c.1525_1527del; NP_000029.2:p.Thr509del | | | | | inframe | | | | **Likely pathogenic** | | | VUS: BP4, PM2_supporting | | | | | |
|  | LOVD APC_000075 | NM_000038.6:c.2546_2551del; NP_000029.2:p.Asp849_Ser851delinsGly | | | | | inframe | | | | **pathogenic** | | | VUS: BP4, PM2_supporting, PS4_moderate | | | | | |
|  | LOVD APC_000883 | NM_000038.6:c.3542_3568del; NP_000029.2:p.Leu1181_Ser1189del | | | | | inframe | | | | **pathogenic** | | | VUS: BP4, PM2_supporting | | | | | |
| *Truncating variants at the 3’ end of the gene and therefore excluded from the application of PVS1* | | | | | | | | | | | | | | | | | | | |
|  | **Database_ID** | **HGVSc; HGVSp** | | | | | **Predicted** | | | **Prior_classification** | | | | | **Reclassification by APC-specific criteria** | | | | |
|  | ClinVar 545875 | NM_000038.6:c.7946_7955del; NP_000029.2:p.Pro2649LeufsTer8 | | | | | frameshift | | | **Pathogenic** | | | | | VUS: PM2_supporting | | | | |
|  | ClinVar 545737 | NM_000038.6:c.7959_7962del; NP_000029.2:p.Thr2654ArgfsTer5 | | | | | frameshift | | | **Pathogenic** | | | | | VUS: PM2_supporting | | | | |
|  | ClinVar 428168 | NM_000038.6:c.8047del; NP_000029.2:p.Ile2683LeufsTer40 | | | | | frameshift | | | **Pathogenic** | | | | | VUS: PM2_supporting | | | | |
|  | ClinVar 827446 | NM_000038.6:c.8099_8102del; NP_000029.2:p.Asn2700ArgfsTer22 | | | | | frameshift | | | **Likely pathogenic** | | | | | VUS: PM2_supporting | | | | |
|  | LOVD APC_001492 | NM_000038.6:c.8344del; NP_000029.2:p.Thr2782LeufsTer28 | | | | | frameshift | | | **pathogenic** | | | | | VUS: PM2_supporting | | | | |
|  | ClinVar, LOVD 486740 | NM_000038.6:c.8514C>A; NP_000029.2:p.Tyr2838Ter | | | | | nonsense | | | **Likely pathogenic** | | | | | VUS: PM2_supporting | | | | |
|  | ClinVar 233392 | NM_000038.6:c.8514C>G; NP_000029.2:p.Tyr2838Ter | | | | | nonsense | | | **Likely pathogenic** | | | | | VUS: PM2_supporting | | | | |
