## Supplemental Table 4 for "Systematic large-scale application of ClinGen InSiGHT *APC*-specific ACMG/AMP variant classification criteria substantially alleviates the burden of variants of uncertain significance in ClinVar and LOVD databases"

**Supplementary Table 4 Frequency of application of the *APC-specific* variant classification criteria**

| **Fulfilled code** | **No. of variants**  **(% of all 10,228 variants)** | **Variants included** |
| --- | --- | --- |
| PM2_supporting | 7083 (69%) | Variants absent from population databases |
| BP1 | 4220 (41%) | Missense variants outside of 1^st^ 15-amino-acid repeat |
| BP4 | 2789 (27%) | Variants with consistently benign splice prediction |
| BP7 | 2236 (22%) | Synonymous, intronic variant without splice effect |
| PVS1_variable | 2192 (21%) | Frameshift, nonsense, spliceogenic, gross deletions and duplications, last nucleotideG to non-G |
| BS1_variable | 2089 (20%) | Variants with allele frequency above BS1 threshold for *APC* |
| PS4_variable | 455 (4%) | Variants present in individuals with scorable phenotypic information |
| BA1 | 427 (4%) | Variants with allele frequency above BA1 threshold for *APC* |
| BS2_variable | 155 (2%) | Variants present in healthy unaffected adult individual |
| PP3 | 67 (1%) | Variants with consistently pathogenic splice prediction |
| PS3_variable | 51 | VCEP-approved functional studies suggestive of a pathogenic effect |
| PS1_variable | 44 | Variants at the same position (same amino acid change) as previously established pathogenic variants |
| PS2_variable | 16 | Confirmed *de novo* variants |
| PM6_variable | 15 | Presumed *de novo* variants |
| BS3_variable | 10 | VCEP-approved functional studies suggestive of a benign effect |
| PP1_variable | 8 | Variants that fulfilled segregation requirements |
| BP5 | 8 | Variants present in individuals with alternative cause of disease |
| PM5_variable | 5 | Variants at the same position (different amino acid change) as previously established pathogenic variants |
| BS4_variable | 2 | Variants that demonstrated a lack of segregation with disease |
| BP2 | 1 | Co-occurrence with established pathogenic *APC* variants |
